# Beliefs about Mask Efficacy and the Effect of Social Norms on Mask Wearing Intentions for COVID-19 Risk Reduction

**DOI:** 10.1101/2021.03.02.21252722

**Authors:** Scott E. Bokemper, Maria Cucciniello, Tiziano Rotesi, Paolo Pin, Amyn A. Malik, Kathryn Willebrand, Elliott E. Paintsil, Saad B. Omer, Gregory A. Huber, Alessia Melegaro

**Author notes:** **Corresponding Author:** Gregory A. Huber, **Email:**. **Author Contributions:** All authors designed the research and contributed to the preparation of the manuscript. SEB, MC, GAH, and AM programmed survey instruments. SEB, MC, TR, GAH AM and PP analyzed the data. **Competing Interest Statement:** The authors declare no competing interests regarding this work.

## Abstract

In the absence of widespread vaccination for COVID-19, governments and public health officials have advocated for the public to wear masks during the pandemic. The decision to wear a mask in public is likely affected by both beliefs about its efficacy and the prevalence of the behavior. Greater mask use in the community may encourage others to follow this norm, but it also creates an incentive for individuals to free ride on the protection afforded to them by others. We report the results of two vignette-based experiments conducted in the United States and Italy to examine the causal relationship between beliefs, social norms, and reported intentions to engage in mask promoting behavior. We find that providing factual information about how masks protect others increases the likelihood that someone would wear a mask or encourage others to do so in the United States, but not in Italy. There is no effect of providing information about how masks protect the wearer in either country. Additionally, greater mask use increases intentions to wear a mask and encourage someone else to wear theirs properly in both the United States and Italy. Thus, community mask use may be self-reinforcing.

## Introduction

The COVID-19 pandemic has produced more than 77 million cases resulting in at least 1.6 million deaths globally as of December 14, 2020.(1) To slow the spread of the virus, governments and public health officials have encouraged, and in many instances mandated, the use of masks or other face coverings in public spaces. The effectiveness of this strategy is determined by the prevalence of mask wearing in the population.(2) Recent research finds that wearing a mask may protect both the wearer and those around them from contracting COVID-19.(3-8) Additionally, people who believe that masks are effective report that they are more likely to wear them.(9)

But, persuasive communication experiments have found mixed positive (10-13) and null effects (14, 15) on whether messaging increases people’s intentions to wear a mask. Several experiments that have found positive effects of persuasive health communication have identified that appealing to protecting others may be a particularly effective strategy. For example, a message that emphasized how COVID-19 is a threat to “your community” increased people’s intentions to wear a mask. (12) Similarly, other work has found that inducing empathy for an individual who is particularly vulnerable to COVID-19 increased mask wearing intentions relative to an information only condition and an untreated control condition. (13) Notably, the information only condition did not increase individuals’ intentions to wear a mask compared to the untreated control. This suggests that purely informational messages may not be effective at changing behavior, though other work has found that an infographic that provides information about how masks slow the spread of COVID-19 changes beliefs about mask wearing. (16)

Increasing mask use is an important public health strategy as the more people who wear masks while in public reduces the spread of the disease. Whether any given individual chooses to wear a mask is likely a function of the degree to which they believe that wearing a mask protects them or those around them. But, given that mask wearing is easily observable, the behavior of others is also likely to influence an individual’s decision to wear a mask. As more people make the decision to wear a mask, it creates an incentive for individuals to free ride on the protection afforded to them by others. This could cause high mask compliance to unravel, thus making everyone less safe. Alternatively, a higher prevalence of mask use creates a stronger social norm that could induce even higher levels of compliance within the population.

Recent survey evidence from Germany suggests that people who wear a mask view others who also do so more favorably than those who do not, which supports the idea that wearing a mask in public is a social norm.(17) Further, people who report that their friends and family wear masks at a greater rate are more likely to do so themselves.(18) This suggests that the decision to wear a mask may be influenced by the behavior of others. However, given the novelty of mask wearing in public places, particularly in the United States and Italy, it is not clear ex ante whether the effects of social norms will outweigh the incentives to free ride.

We conducted two vignette-based survey experiments in two of the countries most affected by COVID-19, one in the United States and one in Italy, to better understand the causal relationship between 1) beliefs about mask efficacy, and 2) others’ masking behavior and mask-relevant behaviors and attitudes. First, we examined whether experimentally inducing changes in beliefs about mask efficacy affects people’s willingness to use them, ask others to use them properly, and judgment of those who do not. We separately examine beliefs about whether masks protect the wearer from sick people or whether masks protect one from spreading the disease to others. Second, we examined how experimentally manipulated prevalence of mask use changes these outcomes.

One experiment was fielded in the United States, which does not have a national mask mandate and where public support for mask wearing is highly variable. The other experiment was fielded in Italy, which prior to our experiment being fielded instituted a national outdoor face mask mandate in an effort to contain the spread of SARS-CoV-2 infection. Respondents in Italy were significantly more likely than those in the United States to report both wearing a mask themselves and observing greater mask use by others in their daily lives (Table 1). Thus, these experiments also allow us to ascertain whether the effect of our experimental treatments are robust across contexts that vary considerably in baseline mask use.

**Table 1.** For own mask use and the observation of others mask use in daily life, Italians report that they are more likely to wear a mask and that they observe more mask wearing around them compared to Americans. Italian respondents and American respondents had similar baseline beliefs about the self-protecting effect of wearing a mask. We find suggestive evidence that Italian respondents were more likely to believe that masks also protect others in the control condition, although this difference was not significant at the conventional 5% level. Outcomes in the table are scaled to range from 0 to 1.

|  | United States<br>Mean (S.D.) | Italy<br>Mean (S.D.) | T statistic (p) |
| --- | --- | --- | --- |
| Own Mask Use | 0.77 (0.31) | 0.89 (0.22) | 17.33 (< .001) |
| Mask Use Around You | 0.58 (0.32) | 0.74 (0.26) | 20.25 (< .001) |
| Masks Protect You (Control) | 0.74 (0.29) | 0.73 (0.28) | 1.10 (.31) |
| Masks Protect Others (Control) | 0.80 (0.26) | 0.82 (0.23) | 1.91 (.06) |

### Design

We asked respondents to read three vignettes and report their own intended behavior or their evaluation of the behavior of a third party.^*^ Our core randomized treatments allow us to test whether beliefs about mask efficacy and the mask wearing behavior of others affect one’s own mask wearing and one’s willingness to encourage mask wearing by others. At the subject level, we randomized respondents at equal rates to a *Placebo Control*, where they read a message unrelated to the topic of the experiment, a *Masks Protect You* treatment that explains how masks stop some percentage of SARS-CoV-2 virus particles in the air from being inhaled by healthy people, and a *Masks Protect Others* treatment that explains how masks stop some percentage of SARS-CoV-2 virus particles from being expelled into the air by sick people. This randomization allowed us to understand if manipulating different beliefs about how masks work has distinct effects on intended behaviors.

At the vignette level, we randomized the behavior of the other people described in the vignette to be either that all/almost all other people were wearing masks or none/very few people were wearing masks. This randomization allowed us to test whether the behavior of others caused free riding in situations involving masks or if high compliance with the social norm of wearing a mask increases the likelihood that an individual also complies with the norm.

The complete experiment proceeded as follows. After providing informed consent, respondents completed a variety of items asked as pre-treatment covariates. Subjects then read their assigned mask efficacy treatment and answered a pair of questions about mask efficacy, described in greater detail below. This allowed us to assess the efficacy of the treatments in affecting beliefs about the degree to which masks are self-protecting or other-regarding. Next, respondents read three experimentally manipulated vignettes and provided their response to each vignette.

Specifically, each respondent completed one each of a scenario about their a) OWN masking behavior, b) behavior toward OTHERS who are not properly wearing a mask, and c) evaluation of a THIRD PARTY who was not wearing their mask properly and someone who took an action in response to that. These evaluations were embedded in vignette setting about a) withdrawing money from an ATM, b) taking a walk in a public PARK, and c) going to a house MEETING in their neighborhood (see SI S-1 for complete details). Each respondent read one scenario in each of the three setting in a random order with each scenario and setting drawn without replacement. These randomizations and the flow of the experiment are summarized in SI S-3.

In the OWN behavior scenarios, respondents were asked what they would do in a situation where they had forgotten a mask. In the OTHER behavior scenarios they were asked what they would do if someone else was not wearing their mask properly. Finally, in the THIRD PARTY scenario, respondents read a scenario as an unaffected third party and rated the behavior of others. Specifically, respondents were asked about their evaluation of someone who is not properly wearing a mask and their evaluation of a person who took an action in response to that, as well as what they believe the person who is not wearing a mask would think about the action that the other person took. In this third party scenario, the behavior of the person toward the third party was randomly assigned from 1) proceeding with the activity as usual, 2) proceeding with the activity, but keeping their distance from the person who was not wearing a mask, 3) stopping the activity, or 4) asking the person to fix their mask.^†^

This design as well as our outcome measurement were pre-registered. For ease of presentation, we present analysis on pooled data by scenario type and discuss the dichotomized outcome measures in the main text. We pre-registered analysis by specific vignette, e.g. ATM, and report that analysis in the SI S-4. Substantive conclusions are the same.

## Results

### Effect of providing information about mask efficacy

We first examine whether providing information about how masks work affects beliefs about the efficacy of masks. Comparing across countries, we observe similar levels of the belief that masks protect you in the control condition, but we find suggestive evidence that Italians may be more likely to believe that masks protect others than Americans at baseline, though this difference was not significant at conventional levels (Table 1). For both countries, we validate that providing information changes beliefs about mask efficacy (see SI Figure S1). Specifically, we observe that providing information about how masks protect the wearer increases beliefs that masks protect you without changing beliefs about whether masks protect others. Similarly, providing information that masks protect others increase beliefs that masks protect others although this effect is not statistically significant in the Italian sample. At the same time, information about how masks protect others also changes beliefs that masks protect the wearer. In practice, this means that we can perturb both sets of beliefs with information about masks protecting others, but can only change beliefs about masks protecting the wearer with that information.

Figure 1 panel A plots the effect of the experimental interventions on reported behavior in the OWN behavior scenario pooling across settings. Panel B plots the same results in the OTHER behavior scenario. Panel A provides clear evidence that providing information about how masks protect the wearer has no effect on own mask wearing behavior, whether measured using a scale (1= Continue activity without a mask, 4=go get a mask) or a binary outcome (1=go get a mask, 0=all other behavior). By contrast, providing information about masks protects others increases reported risk reduction behavior in the US but not in Italy. For example, in the US sample, focusing on a dichotomous measure of mask wearing, the masks protect others intervention increases willingness to get a mask by 5.6 percentage points (95% C.I. = 1.4% to 9.7%, *p* < .01), an increase of 10.3% from the baseline level of 54.2% in the control group. (Notably, we can likely attribute these effects to changes in beliefs about protecting others, because this is the only belief perturbed by the Masks Protect Others treatment that is not also affected by the Masks Protect You message.) These effects are similar in magnitude among respondents who saw the OWN behavior scenario immediately after the mask efficacy treatment (see Supplementary Materials). Examining by scenario, the effect of the Masks Protect Others treatment was largest in the MEETING scenario, 6.8 percentage points (95% C.I. = 0.05% to 13.5%, *p* < .05), similar in magnitude to the pooled effect, though imprecise, for the ATM scenario, 5.3 percentage points (95% C.I. = −2.3% to 12.9%, *p* = .17), and smallest in the PARK scenario, 2.0 percentage points (95% C.I. = −5.3 % to 9.3%, *p* = .58; See Supplementary Materials). In the Italian sample, the point estimates across all measures are negative, close to zero, and not statistically significant.

**Figure 1.**
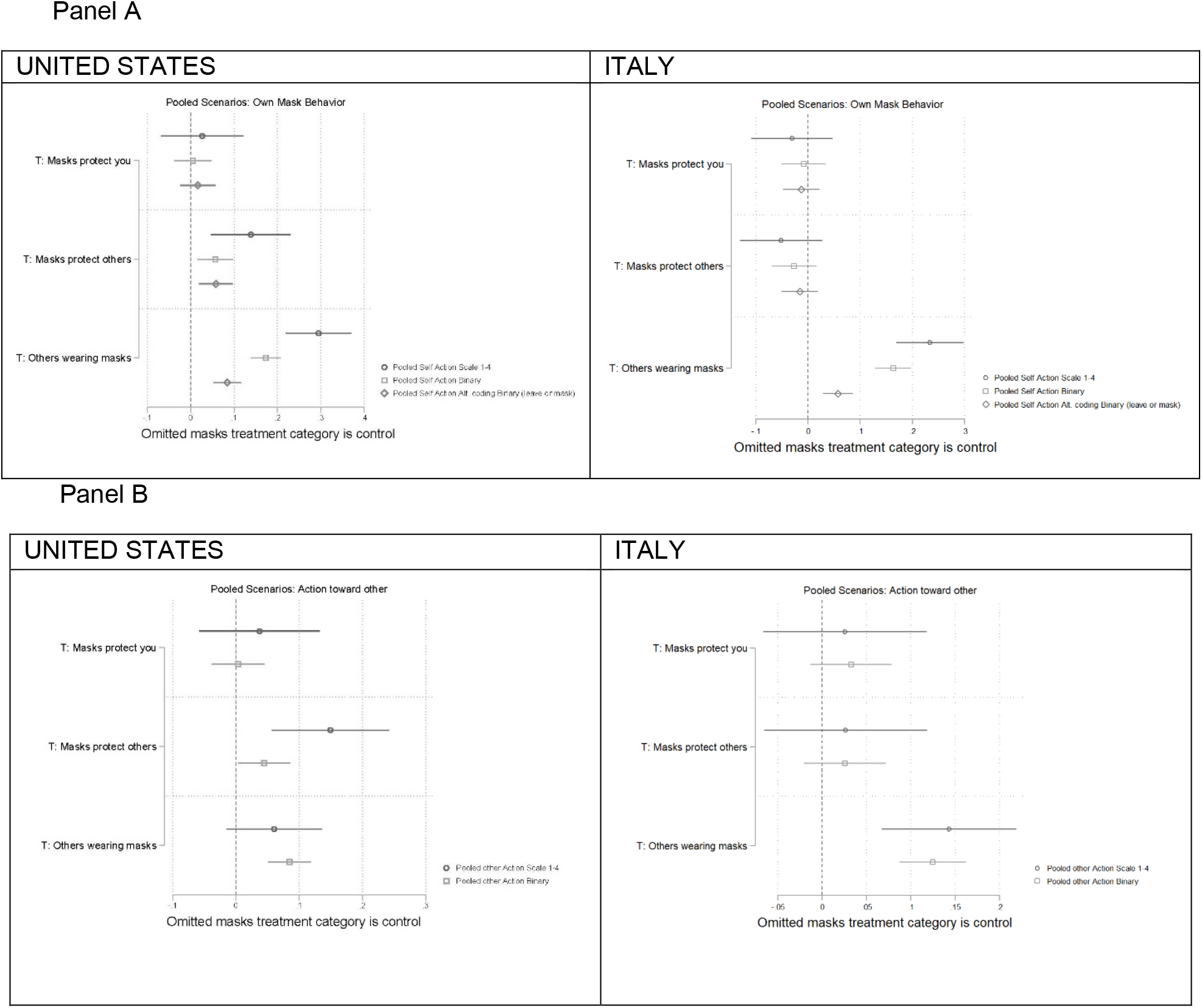
Panel A shows the effect of the mask efficacy treatments and social norms treatment for the OWN mask behavior outcomes. Panel B shows the same treatment effects for the OTHERS outcomes. The figure displays OLS regression estimates with 95% confidence intervals. Models include covariates described in SI S-2. For the United States, we observe a positive and statistically significant effect of the Masks Protect Others treatment on the OWN and OTHERS outcomes. For the others wearing masks treatment, participants in both the United States and Italy were more likely to report that they would retrieve their mask or ask someone else to fix their mask when most or all of the other people described in the vignette were wearing their mask.

Results are similar in the Panel B plot where the outcome is behavior toward the person who is not correctly wearing a mask: Providing information about how masks protect you has small and statistically insignificant effects, while providing information about how masks protect others increases both outcomes in the US but not it Italy. In the US, for the dichotomous willingness to ask another person to fix their mask the effect is 4.4 percentage points (95% C.I. = 0.3% to 8.5%, *p* < .05), an increase of 13.6% from the baseline level of 32.5% in the control group. The estimates are less precise, but similar in magnitude, among respondents who saw the OTHER behavior scenario immediately after the mask efficacy treatment and at the scenario level (see Supplementary Materials). The effect is again small and statistically insignificant in the Italian sample.

We display results from the third-party judgment scenario in Figure 2. Panel A plots evaluations of the person who is not correctly wearing their mask, while Panel B plots evaluations of the person who asks that person to fix their mask. Per Panel A, in neither sample is there an effect of either mask efficacy treatment on the judgment of the person incorrectly wearing their mask. Additionally, per Panel B, across countries, neither treatment has a statistically significant effect on judgments of the person who asks the person to fix their mask.

**Figure 2.**
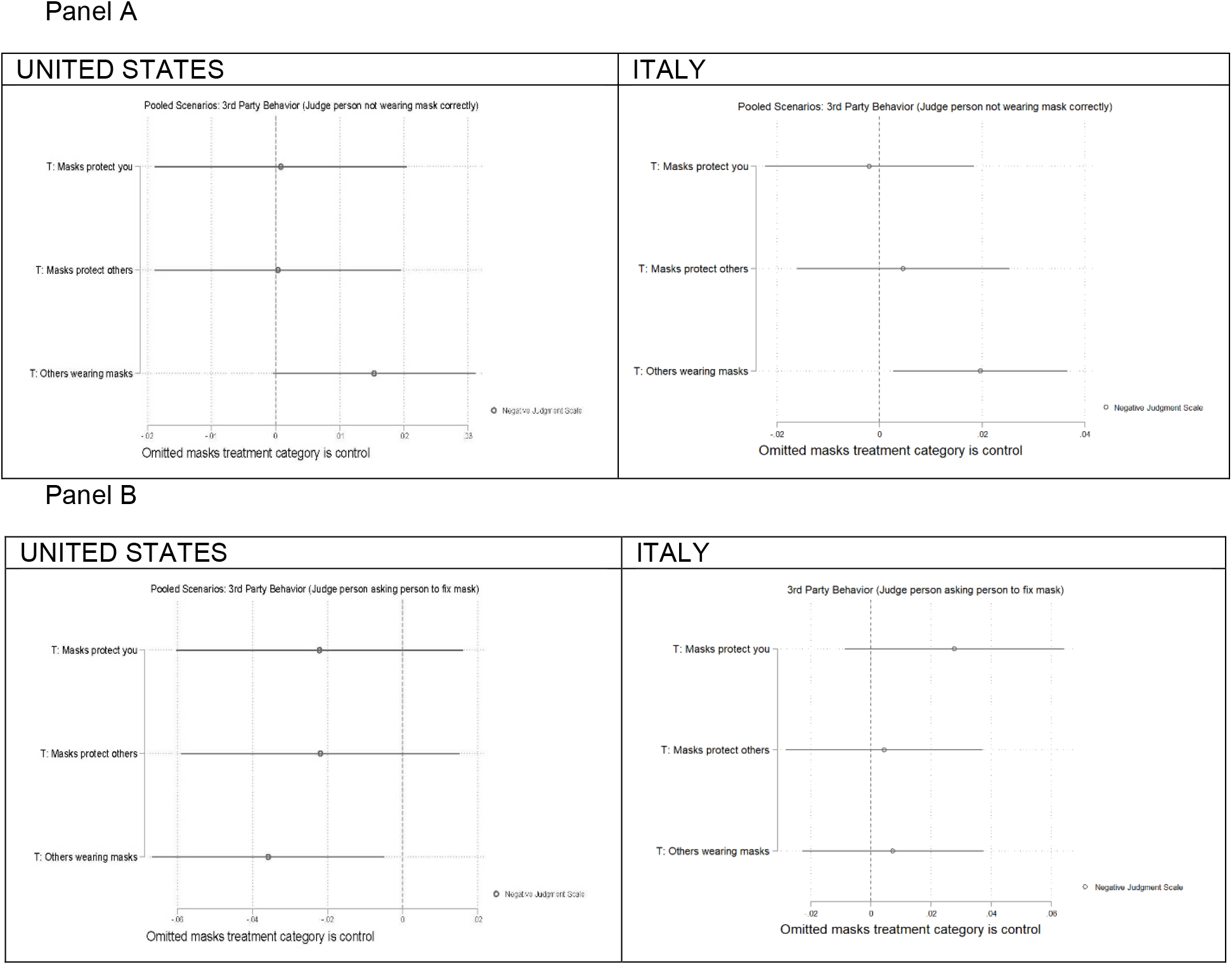
Panel A shows the effect of the mask efficacy treatments and social norms treatment for judging someone who was not wearing their mask appropriate in the THIRD PARTY scenario. Panel B shows the effect of these treatments for judging someone who asked a person to fix their mask in the THIRD PARTY scenario. The figure displays OLS regression estimates with 95% confidence intervals. Models include covariates described in SI S-2. Italian participants were more likely to judge negatively someone who was not wearing their mask properly when everyone else was wearing their mask properly, while participants in the United States viewed someone who asked another person to fix their mask more positively when everyone else was wearing their mask properly. The mask efficacy treatments did not have statistically significant effects on these outcomes.

### Effect of providing information about others’ behavior

We next turn to examining the effect of the behavior of other people described in the vignettes, specifically whether others were wearing masks properly. As a reminder, it is unclear whether more prevalent mask wearing will increase or decrease others’ willingness to wear masks or push others to do so. If others are wearing masks, the risks of contracting COVID-19 are already reduced and people may not wear a mask because they receive protection from others. Additionally, the more people who are wearing masks, the more people who could intervene in the situation. Alternatively, if social norm effects are larger, others wearing masks could cause greater mask wearing and enhance individuals’ willingness to act toward others.

Overall, the evidence strongly leads toward the latter norms interpretation. Returning to Panel A of Figure 1, the effect of other people wearing masks rather than not is to increase the likelihood someone returns to get their own mask by 17.3 percentage points (95% C.I. = 13.9% to 20.8%, *p* < .01) and 16.3 percentage points (95% C.I. = 12.8% to 19.8%) for, respectively, the US and Italy, increases of 32% and 27% compared to baseline. Per Panel B, it also increases the willingness to ask others to fix their mask by 8.4 percentage points (95% C.I. = 5.1% to 11.8%, *p* < .01) in the US and 12.5 percentage points (95% C.I. = 8.7% to 16.2%, *p* < .01) in Italy, increases of 26% and 29.4% compared to baseline levels. Analyzing the data by specific scenario, the effect of other people wearing masks was positive and significant for both one’s willingness to get their own mask and to ask others to fix their mask for all three scenarios in the US and in the ATM and MEETING scenarios in Italy.

This pattern is also apparent in the third-party evaluation scenario. Per panel A of Figure 2, a person who is not wearing their mask correctly is judged more negatively when more other people are wearing their masks, although this effect is not statistically significant in the US (effect of .015 units in US, 95% C.I. = -.001 to .031, *p* = .06; effect of .02 units in Italy, 95% C.I. = .003 to .037, *p* = .02). Similarly, people judge a third party who asks someone else to fix their mask significantly less negatively in the US (B=-.035, 95% C.I. = -.005 to -.067, *p* < .05) when others are wearing their masks, with a null effect in the Italian sample.

Overall, in describing their own masking behavior, their behavior toward others, and their evaluation of third parties, the normative effect of others masking behavior on outcomes is clear across all of these scenarios in the US, but less clear in the Italian cases. Thus, despite concerns that more ubiquitous mask wearing might undercut further encouragement of masking, the effects are the opposite: Compliance by others appears to enhance both individuals’ own behavior and support for behavior towards others. Importantly, there are no cases in which more frequent mask wearing depress others’ willingness to wear masks or ask others to properly wear their masks.

## Discussion

Persuading individuals to engage in COVID-19 risk reduction is an important problem of public policy and public health. We demonstrate that in trying to encourage greater mask wearing, interventions that increase the belief that masks protect others appear promising, particularly in the United States. This suggests that factual information about how facemask use protect others are more effective than messages that emphasize how masks protect the wearer for increasing facemask use and encouraging others to do so. These effects are stronger in the US than in Italy, which may indicate that the small differences in beliefs at baseline were consequential or differences in how each sample is persuaded by these messages. Similarly, messages communicating social benefits and providing prosocial nudges have been shown to increase vaccination intentions in other studies.(19, 20) These social nudges have also been found to be effective in other contexts.(21)

Additionally, we show that increased mask wearing may create self-reinforcing cycles that further promote the behavior. Increased mask wearing does not lead to free riding behavior or a willingness to defer to others in enforcing mask wearing. Instead, more frequent mask wearing creates a stronger social norm inducing better compliance with the mask wearing behavior in both the US and Italy. These findings are robust to various scenarios and vignettes tested in our experiment. Our results are consistent with those by Betsch et al.(19) and McKillop et al.(22) showing that emphasizing the high levels of neighborhood vaccination and the social benefits of vaccination did not induce free riding behavior. Other work has shown that peer influence can inform vaccination decisions, with these effects increasing as overall vaccination rates increase.(23) Our study suggests similar dynamics in an environment where peer influence is perhaps even more important, private mask-wearing behavior.

Our results were consistent in two independently conducted studies in two different settings: the US and Italy. This is important as the US did not have a facemask mandate while Italy did, increasing the generalizability of our results (although beliefs were harder to change in the Italian case). But both the US and Italy are high income western countries. Hence our findings need to be evaluated in other societies for applicability. However, previous work has shown that different cultures are receptive to prosocial messaging leading to increased intentions to perform the desired action, especially in individualistic societies.(20)

We note three limitations of this study. First, our measures are not behavior, but are instead measures of behavioral intentions. The evidence presented here should be used for the design of field trials to validate these estimates in the non-survey setting and with broader populations. Second, our third-party enforcement scenarios involve situations where someone has a mask but is not wearing it correctly, a less difficult case than when someone confronts a third party who does not have a mask at all. Last, we have not investigated how these effects are likely to change as vaccination against COVID-19 becomes more prevalent. It may be that the possibility someone is vaccinated will decrease the willingness to try to convince others to wear a mask because of ambiguity about whether another person is a risk factor. These questions are ripe for further investigation in follow-up work.

Our findings suggest first that public health communication campaigns that emphasize messages about how facemask use protect others are more likely to be effective in promoting mask wearing than campaigns that highlight how facemasks protect the wearer. The greater efficacy of this other-regarding message is clear in the United States, but this may not apply in other contexts as demonstrated by the results in Italy where neither mask efficacy treatment was effective.

Additionally, we find that increasing mask use increases the willingness of others to wear masks and to encourage others to do so also, reducing concerns about how more prevalent mask wearing will lead to free-riding. The magnitudes of the effects of peer behavior are large and robust in both the American and the Italian context.

## Materials and Methods

Both experiments were fielded using samples provided by the survey vendor Lucid. Subjects completed the survey online. The survey was programmed and data collected using the survey software Qualtrics. The United States experiment was fielded between October 1 and October 21, 2020 with a total sample of 3,100 respondents. The Italy experiment was fielded between October 22, 2020 and November 8, 2020 with a total sample of 2,659 respondents.

The survey started with an unrelated vignette used to assess respondent attentiveness. Respondents who did not pass a comprehension question exited the sample. Next, respondents answered questions about their experience with COVID-19 followed by basic demographic questions. Then, respondents read the mask efficacy treatment that they were assigned to and could not advance the page for at least 20 seconds to give them adequate time to read it. Following this, they reported their beliefs about who masks protect and then proceeded to the vignettes that were presented in a random order.

The mask efficacy treatment text and experimental vignettes for both experiments can be found in SI S-1. We estimate treatment effects using OLS regression with robust Huber-White standard errors independently for each experiment. Regressions included controls for settings and demographic covariates that are specified in SI S-2.

## Data Availability

Data will be made available upon publication of the manuscript

## Ethics Statement

The experiments reported here were approved by an IRB in the United States and an IRB in Italy. Informed consent was obtained before participants started the study and subjects were informed that they could terminate their participation at any time. All data was collected anonymously.

## Data availability

Materials to replicate the results presented here will be posted in a public data depository upon publication.

## Acknowledgments

SEB and GAH acknowledge support from the Institution for Social and Policy Studies, the Center for the Study of American Politics, and the Tobin Center for Economic Policy at Yale University. PP and AM acknowledge support from the Italian Ministry of Education Progetti di Rilevante Interesse Nazionale (PRIN), respectively grant number 2017ELHNNJ and 20177BRJXS. AAM, SBO, EP, and KW were supported by Yale Institute for Global Health.

## Footnotes

* In two pilot experiments conducted in the United States and in Italy to pretest the realism of the vignettes, participants, on average, reported that the vignettes were mostly realistic.

† In the third party scenario, we also randomize the names and genders of the individuals involved to allow us to check for the robustness of effects across name and gender combinations.

