## Supplementary Materials for "Beliefs about Mask Efficacy and the Effect of Social Norms on Mask Wearing Intentions for COVID-19 Risk Reduction"

**This PDF file includes:**

S-1 Mask Efficacy Treatments and Vignettes
S-2 Covariates for Analyses and Measures of Mask Wearing
S-3 Progression of Experiment and Randomizations
S-4 Attrition
S-5 Figs. S1 to S15
S-6 Tables S1 to S3
S-7 References

### S-1 Mask Efficacy Treatments and Vignettes

#### Mask Efficacy Treatments

*Placebo Control*

What are the costs and benefits of bird feeding?

It is difficult to assess the costs and benefits of bird feeding because it is difficult to compare the health of birds without access to feeders with birds that are frequent feeders. Only one study was able to obtain some sound results. That study found that any benefits of feeding only appear to occur sporadically under extreme climactic conditions. No research has been able to demonstrate a cost. Aside from costs and benefits to birds, there is a cost of benefit to humanity. The costs are obvious—the expense of bird feeding supplies.

The benefits include learning more about birds and the joy of connecting with the natural world. Bird feeding provides a direct, intimate view of the natural world for more than 50 million Americans who feed the birds in their yards. It is most popular in winter, when birds seem to need the most help. Some people worry that birds will suffer unless they make great efforts to keep the feeder filled, but research indicates that most birds do not depend on feeders.

*Masks Protect You*

Scientists have shown that wearing a face covering over your mouth and nose substantially reduces your risk of COVID-19 infection by decreasing the amount of virus that you inhale into your nasal passages and lungs.

When a sick individual breathes, sneezes, or coughs, virus particles are expelled into the air where they can be inhaled by others. The more of the virus that makes it into your body, the greater your chances of getting sick with COVID-19.

Scientists tested masks to see if they would stop virus particles from getting into a person's body. They found that an N95 respirator stops at least 95% of virus particles, while a simple cotton cloth mask covering your nose and mouth stops approximately 55% of virus particles. (1-3)

*Masks Protect Others*

Scientists have shown that wearing a face covering over your mouth and nose substantially reduces your risk of spreading COVID-19 to others by decreasing the amount of virus that makes it into the air where it can infect other people.

When a sick individual breathes, sneezes, or coughs, virus particles are expelled into the air where they can be inhaled by others. The more of the virus that makes it into someone's body, the greater their chances of getting sick with COVID-19.

Scientists tested masks to see if they would stop virus particles from getting into the air around them. They found that an N95 respirator stops at least 85% of virus particles, while a simple cotton cloth mask covering your nose and mouth stops approximately 80% of virus particles. (2, 4)

*Mask Outcomes*

How much do you agree or disagree with the following statements about how masks might work to reduce the spread of COVID-19? (Strongly disagree, Somewhat disagree, Neither agree nor disagree, Somewhat agree, Strongly agree)

- A mask reduces the risk that sick people spread COVID-19 into the air around them
- A mask reduces the risk that healthy people get COVID-19 from the air around them

#### Vignettes

**Authors coding note:** For the OWN behavior and OTHERS behavior vignettes, the figures in the main text display outcomes estimated three different ways. The first assigns higher values to behaviors that do more to reduce the spread of COVID-19 ranges from 0 for continue the activity as normal, .333 for continue the activity but distance, .667 as stopping the activity, and 1 for taking an action that promotes mask wearing, either for yourself or for someone else reports estimated effects. The second is a dichotomous coding of the variable that takes the value of 1 for mask wearing and 0 otherwise. The third assigns a value of 1 for mask wearing or leaving and a value of 0 otherwise. The first and second coding of the outcomes were pre-registered.

ATM Vignettes

*Own Behavior Vignette*

Suppose you have to go to the ATM machine to withdraw some money. You arrive at the machine and there is a line of 2 people in front of you waiting to use the outdoor machine. [*Social Norms Manipulation*: Both people in line are wearing their masks properly OR Neither person in line is wearing their mask properly].

You realize that you have left your mask at home which is about a 5 minute walk away.

What do you do?

- Continue to wait in line because the risk is low
- Continue to wait in line, but make sure to keep your distance from other people (coded
- Go back home to get your mask and then come back to the ATM
- Give up on getting money from the ATM

*Others Behavior Vignette*

Suppose you have to go to the ATM machine to withdraw some money. You arrive at the machine and there is a line of 2 people in front of you waiting to use the outdoor machine. [*Social Norms Manipulation*: Both people in line are wearing their masks properly OR Neither person in line is wearing their mask properly].

Someone you know joins the line behind you. They have a face mask, but it is hanging around their neck and not covering their nose or mouth.

What do you do?

- Continue to wait in line because the risk is low
- Continue to wait in line, but make sure to keep your distance from the new person because they aren't wearing their mask properly
- Ask the person behind you to their mask on over their mouth/nose
- Leave the line and come back when the person who isn't wearing their mask properly is gone

*Third Party Vignette*

Suppose [Erin/Chris] goes to the ATM machine to withdraw some money. [He/She] arrives at the machine and there is a line of 2 people in front of [him/her] waiting to use the outdoor machine. [*Social Norms Manipulation*: Both people in line are wearing their masks properly OR Neither person in line is wearing their mask properly].

[Chris/Erin], someone who belongs to the same gym as [Erin/Chris], joins the line behind [Erin/Chris]. [Chris/Erin] has a face mask, but it is hanging around [his/her] neck and not covering [his/her] nose or mouth.

Based on what you've read so far, how much do you think each of these words describes [Chris/Erin]? (0 Not at all, .25 Slightly, .5 Somewhat, .75 Mostly, 1 Very)

- Intelligent
- Trustworthy
- Selfish
- Competent
- Aggressive
- Likeable
- Reckless

[Erin/Chris] notices how [Chris/Erin] is wearing [his/her] mask. [Erin/Chris] decides to [1. continue waiting in line because the risk is low OR 2. continue waiting in line, but make sure to keep [his/her] distance from [Chris/Erin] because [Chris/Erin] isn't properly wearing a mask OR 3. ask [Chris/Erin] to put [his/her] mask over [his/her] mouth and nose OR 4. leave the line and come back when [Chris/Erin] is gone].

Based on what you've read so far, how much do you think each of these words describe [Chris/Erin]? (0 Not at all, .25 Slightly, .5 Somewhat, .75 Mostly, 1 Very)

- Intelligent

- Trustworthy
- Selfish
- Competent
- Aggressive
- Likeable
- Reckless

PARK Vignettes

*Own Behavior Vignette*

Suppose you have driven to a local park to go for a 2 hour walk this weekend. The park is crowded with many other people and families out enjoying the good weather.

After parking your car, you walk about 5 minutes into the main area of the park. You suddenly realize that you have left your own face mask in your car. [*Social Norms Manipulation*: Not many of the other people at the park are properly wearing a face mask OR Almost all of the other people at the park are properly wearing a face mask].

What do you do?

- Continue your walk because you are outside and the risk of anything bad happening when you're outside is small
- Continue your walk, but make sure to avoid other people because you do not have a mask
- Return to your car to retrieve your mask
- Return to your car and leave the park

*Others Behavior Vignette*

Suppose you have driven to a local park to go for a 2 hour walk this weekend. The park is crowded with many other people and families out enjoying the good weather.

After parking your car, you put on your face mask and walk about 5 minutes into the main area of the park. [*Social Norms Manipulation*: Not many of the other people at the park are properly wearing a face mask OR Almost all of the other people at the park are properly wearing a face mask].

You run into someone from your neighborhood, who asks if they can join you for your walk. However, when they approach you suddenly realize that they have their face mask hanging around their neck, rather than covering their nose and mouth, while you are properly wearing your mask.

What do you do?

- Continue your walk because you are outside and the risk of anything bad happening is small

- Continue your walk, but make sure to stay distant from your acquaintance because they do not have their mask on properly
- Tell your acquaintance that you are about to leave the park and head back to your car
- Ask your acquaintance to put their mask on so it covers their nose and mouth

*Third Party Vignette*

Suppose [Erin/Chris] and [Chris/Erin], two neighbors, run into each other at a local park this weekend. The park is crowded with many other people and families out enjoying the good weather. [*Social Norms Manipulation*: Not many of the other people at the park are properly wearing a face mask OR Almost all of the other people at the park are properly wearing a face mask].

[Erin/Chris] is properly wearing [his/her] face mask so it covers [his/her] nose and mouth, while [Chris/Erin] has [his/her] mask hanging around [his/her] neck, rather than covering [his/her] nose and mouth.

Based on what you've read so far, how much do you think each of these words describes [Chris/Erin]? (0 Not at all, .25 Slightly, .5 Somewhat, .75 Mostly, 1 Very)

- Intelligent
- Trustworthy
- Selfish
- Competent
- Aggressive
- Likeable
- Reckless

[Erin/Chris] notices how [Chris/Erin] is wearing [his/her] mask. [Erin/Chris] decides to [1. continue [his/her] walk because [he/she] is outside and the risk of anything bad happening is small OR 2. continue [his/her] walk, but makes sure to stay distance from [Chris/Erin] because [he/she] does not have [his/her] mask on properly OR 3. tell [Chris/Erin] that [he/she] is about to leave the park and head back to [his/her] car OR 4. ask [Chris/Erin] to put [his/her] mask on properly so that it covers [his/her] nose and mouth].

Based on what you've read so far, how much would you think each of these words describe [Erin/Chris]? (0 Not at all, .25 Slightly, .5 Somewhat, .75 Mostly, 1 Very)

- Intelligent
- Trustworthy
- Selfish
- Competent
- Aggressive
- Likeable
- Reckless

MEETING Vignettes

*Own Behavior Vignette*

Suppose you've been invited by someone living in your neighborhood to a five-person meeting at their house. The meeting will be held in their living room.

You arrive at their house, which is about 5 minutes from your own home. You pull into the driveway to park. You now remember that you didn't bring a mask with you. [*Social Norms Manipulation*: None of the other guests are properly wearing a face mask OR All of the other guests are properly wearing a face mask].

What do you do?

- Go to the meeting because the risk of anything bad happening is small
- Go the meeting, but try to stay away from other people because you do not have a mask
- Send your neighbor a text saying that you've forgotten something and will be right back, and return home to get a mask
- Send your neighbor a text apologizing for having to leave, but say something has suddenly come up.

*Other Behavior Vignette*

Suppose you've been invited by someone living in your neighborhood to a five-person meeting at their house. The meeting will be held in their living room.

You arrive at their house, which is about 5 minutes from your own home. You pull into the driveway to park. [*Social Norms Manipulation*: None of the other guests are properly wearing a face mask OR All of the other guests are properly wearing a face mask]. The three guests who arrived before you go inside while you are parking.

You have gotten out of your car and put on your mask when the last guest arrives. They get out of their car too, but have their mask hanging around their neck, rather than covering their face and mouth.

What do you do?

- Go to the meeting as is because the risk of anything bad happening is small
- Go the meeting, but make sure to stay distant from this person because they do not have their mask on properly
- Ask this guest to put their mask on so it covers their nose and mouth
- Send your neighbor a text apologizing for having to leave, but say that something has suddenly come up so you can't come to the meeting

*Third Party Vignette*

Suppose [Erin/Chris] has been invited by Pat, a neighbor, to a five-person meeting at Pat's house. The meeting will be held in Pat's living room.

[Erin/Chris] arrives at Pat's house, which if about 5 minutes from [Erin/Chris]'s home. [Erin/Chris] pulls into the driveway to park. [*Social Norms Manipulation*: None of the other guests are properly wearing a face mask OR All of the other guests are properly wearing a face mask]. The three guests who arrived before [Erin/Chris] go inside while [his/her] is parking.

[Erin/Chris] has gotten out of [Erin/Chris] car and put on [Erin/Chris] mask when the last guest, [Chris/Erin], arrives. [Chris/Erin] gets out of [his/her] car too, but has [his/her] mask hanging around [his/her] neck, rather than covering [his/her] face and mouth.

Based on what you've read so far, how much do you think each of these words describes [Chris/Erin]? (0 Not at all, .25 Slightly, .5 Somewhat, .75 Mostly, 1 Very)

- Intelligent
- Trustworthy
- Selfish
- Competent
- Aggressive
- Likeable
- Reckless

[Erin/Chris] notices how [Chris/Erin] is wearing [his/her] mask. [Erin/Chris] decides to [1. go to the meeting as is because the risk of anything happening is small OR 2. go to the meeting, but makes sure to stay distant from [Chris/Erin] .

Based on what you've read so far, how much would you think each of these words describe [Erin/Chris]? (0 Not at all, .25 Slightly, .5 Somewhat, .75 Mostly, 1 Very)

- Intelligent
- Trustworthy
- Selfish
- Competent
- Aggressive
- Likeable
- Reckless

### Mask Efficacy Treatments and Vignettes Italian Study

The survey questionnaire treatments and vignettes were translated into Italian by the team of Italian researchers. Some specific elements were adapted to the Italian context to make the survey more real and coherent with the national setting.

The names Erin and Chris were replaced with Sofia and Francesco the two most common names in Italy based on recent data (Istat, 2019).

The text for the placebo control was modified to make it more fitting with the Italian context and the story of the “moka pot” was included (The use of the moka pot vignette was not pre-registered and the change from the bird feeding vignette was made prior to any data collection).

Following translation, the instrument was pilot tested on 180 university students. During this test the respondents were asked specific debriefing questions in order to determine if the survey questions were understood and if the intent of the question was accurately conveyed.
The full Italian survey is available upon request.

#### Mask Efficacy Treatments

*Placebo Control*

La caffettiera moka per caffè è un piccolo apparecchio che serve a fare il caffè in casa, estremamente diffuso in quasi tutte le case d’Italia. Fu una fortunata invenzione di Alfonso Bialetti il cui figlio Renato rese poi famosa in tutto il mondo la caffettiera. Le dimensioni variano a seconda di quanto caffè si desidera ottenere. Il caffè si misura in "tazze" o in "persone" e una moka si definisce pertanto da 1, 2, 3 o piu’ persone o tazze.
La moka è formata da 5 elementi che si montano ad incastro tra loro: la caldaia, munita di valvola di sicurezza; il serbatoio del caffè, a forma di imbuto con piano filtrante, dove si mette la polvere di caffè torrefatto; il filtro che trattiene la polvere del caffè ed evita che resti in sospensione nella bevanda finale; la guarnizione di gomma, che trattiene la piastrina in posizione ed evita la fuoriuscita laterale di acqua e vapore, alla base del serbatoio del caffè; il bricco, che raccoglie la bevanda stessa e che si chiude a vite sul serbatoio dell'acqua. Questo è provvisto di cannula da cui esce il caffè che si è formato per contatto tra l'acqua bollente e la polvere. Il contenitore finale ha un beccuccio dal quale si versa la bevanda ed una maniglia per afferrare l'apparecchio.
La preparazione consiste nel versare l'acqua nella caldaia, tapparla con il filtro che viene riempito di polvere di caffè, chiudere il tutto con il corpo della caffettiera e porlo sul fuoco.L'acqua scaldandosi fa aumentare la pressione all'interno e risale verso l'alto, passando prima nel filtro dell'imbuto, filtrando nella polvere del caffè, e quindi nel filtro della piastrina. Attraverso un beccuccio fuoriesce nel contenitore superiore. Una volta riempito il contenitore superiore, il caffè è pronto per la degustazione.

#### S-2 Covariates for Analyses

##### United States Covariates

*Age* in years

*Gender*, entered as categories (Male, Female, Other/Missing)

*Household Income* (Categories provided by Lucid, with a separate indicator for choosing not to supply income/missing)

*Education*, entered as categories (As provided by Lucid)

*Partisanship* (Indicators for each of the 7 standard partisanship categories, Strong Republican, Weak Republican, Lean Republican, Independent/Other/Missing, Lean Democrat, Weak Democrat, Strong Democrat)

*Race* (Indicators for Black, Latino, and Other Races)

*Employment* (Working at home, working outside home by choice, Working outside home because required, not working)

*Flu shot* In the last 5 years, how many times have you gotten the seasonal flu vaccine (flu shot)? (0, 1, 2, 3, 4, 5)

##### Italy Covariates

*Age* in years

*Gender*, entered as categories (Male, Female, Other/Missing)

*Household Income* (Categories provided by Lucid, with a separate indicator for choosing not to supply income/missing)

*Education*, entered as categories (As provided by Lucid)

*Vote (*Indicator for each of 7 alternatives regarding vote in 2016 elections: Did not vote, Potere al Popolo, Liberi e Uguali, Partito Democratico (coalition), Forza Italia/Lega/Fratelli D'Italia, Casa Pound, Movimento 5 Stelle, Doesn't know).

*Employment* (Working at home, working outside home by choice, Working outside home because required, not working)

*Flu shot* In the last 5 years, how many times have you gotten the seasonal flu vaccine (flu shot)? (0, 1, 2, 3, 4, 5)

**Mask Wearing Items**

*Your Mask Use*
How often do you wear a mask?

- I refuse to wear a mask, even when it is required by a business or law.
- I only wear a mask when I'm required to by a business or law.
- I occasionally wear a mask when I'm out in public, even if it is not required.
- I usually wear a mask when I'm out in public, even if it is not required.
- I always wear a mask when I'm out in public, even if it is not required.

*Others Mask Use*
When you go out in your neighborhood and community, how many people that you see out in public are wearing a mask?

- 0-20% (1 out of 5 or fewer people)
- 21-40% (between 1 out of 5 and 2 out of 5 people)
- 41-60% (about half)
- 61-80% (between 3 out of 5 and 4 out of 5 people)
- 81-100% (more than 4 out of 5 people)

#### S-3 Progression of the Experiment and Randomizations

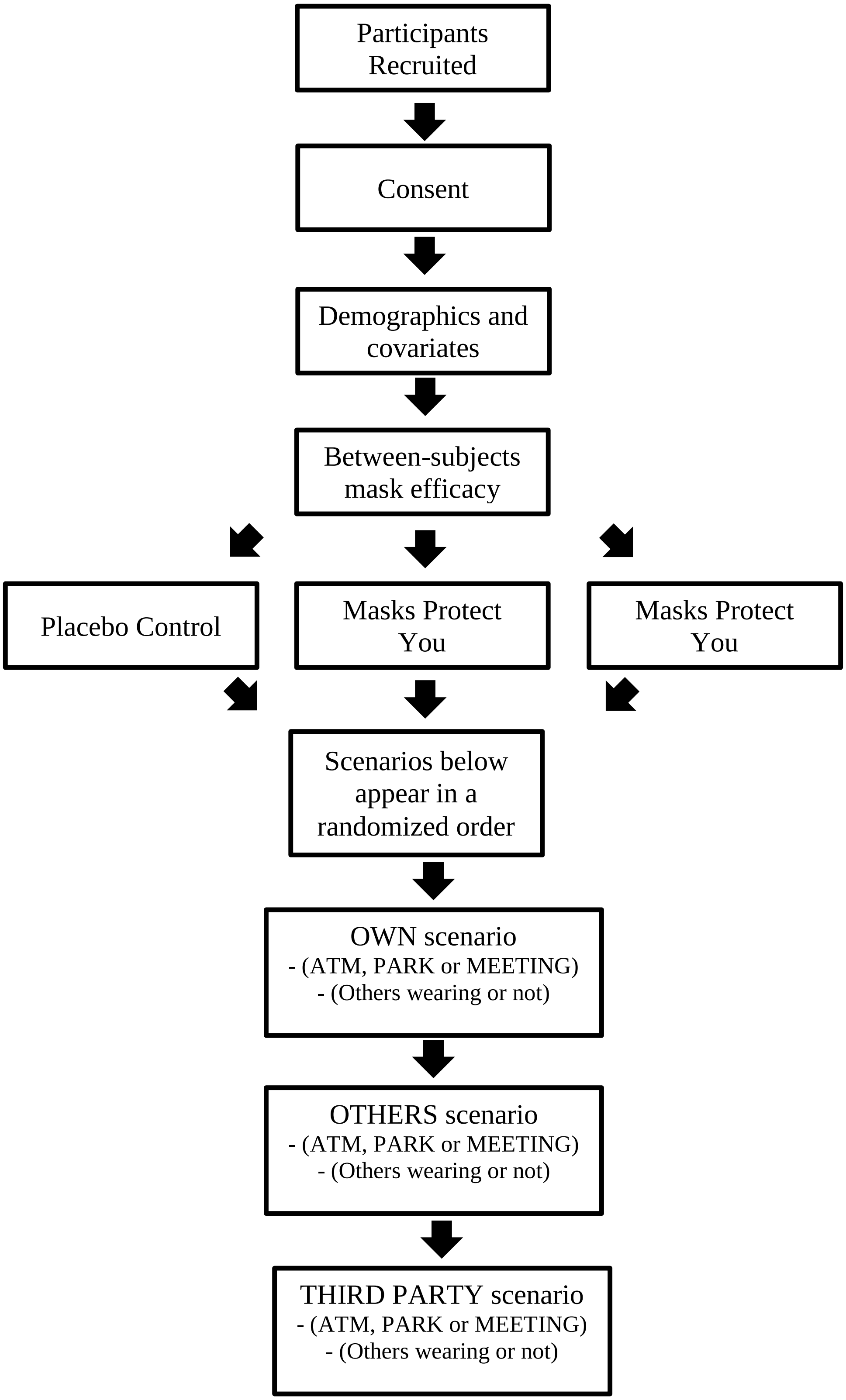

Sample Sizes by Treatment

| Treatment | U.S. Sample Size | Italian Sample Size |
|---|---|---|
| **Mask Efficacy Treatments** | | |
| Untreated Control | 965 | 855 |
| Masks Protect You | 1,023 | 882 |
| Masks Protect Others | 1,112 | 850 |
| **Vignette Treatments** | | |
| ATM Scenario | | |
| OWN | 1,011 | 856 |
| OTHERS | 1,067 | 867 |
| THIRD PARTY | 1,022 | 864 |
| Park Scenario | | |
| OWN | 985 | 868 |
| OTHERS | 1,055 | 870 |
| THIRD PARTY | 1,060 | 849 |
| Meeting Scenario | | |
| OWN | 1,104 | 863 |
| OTHERS | 978 | 850 |
| THIRD PARTY | 1,018 | 874 |
| **Behavior of Others** | | |
| ATM Scenario | | |
| Both people in line are wearing their masks properly | 1,559 | 1,274 |
| Neither person in line is wearing their mask properly | 1,541 | 1,313 |
| Park Scenario | | |
| Almost all of the other people at the park are properly wearing a face mask | 1,528 | 1,294 |
| Not many of the other people at the park are properly wearing a face mask | 1,572 | 1,293 |
| Meeting Scenario | | |
| All of the other guests are properly wearing a face mask | 1,555 | 1,279 |
| None of the other guests are properly wearing a face mask | 1,545 | 1,308 |

*Attrition by Condition*

We examine within study attrition as defined by respondents who answered the question immediately prior to viewing the mask efficacy treatment. For the experiment in the United States, we had a total of 74 participants or 2.3% of the sample that were exposed to the treatment drop out of the study prior to completing any of the vignettes (17 in the control, 28 in the Masks Protect You treatment, and 29 in the Masks Protect Others treatment). We used OLS regression with Huber-White standard errors to predict attrition by condition. For the Masks Protect You and the Masks Protect Others condition, we did not find that any of the covariates detailed above predicted attrition. Shifting to the control condition, we find some evidence that political independents were more likely to drop out. Given the small amount of attrition in this condition, it is unlikely that it biases the treatment effects.

For the Italian sample, 51 participants or 2% of the sample drop out of the study prior to completing any of the vignettes (19 in the control, 18 in the Masks Protect You treatment, and 14 in the Masks Protect Others treatment). Somewhat in line with the US sample, we find that people who do not identify with any of the major Italian parties are more likely to drop out.

### S-4 Supplementary Figures

#### Figure S1

| United States | ITALY |
|---|---|
| 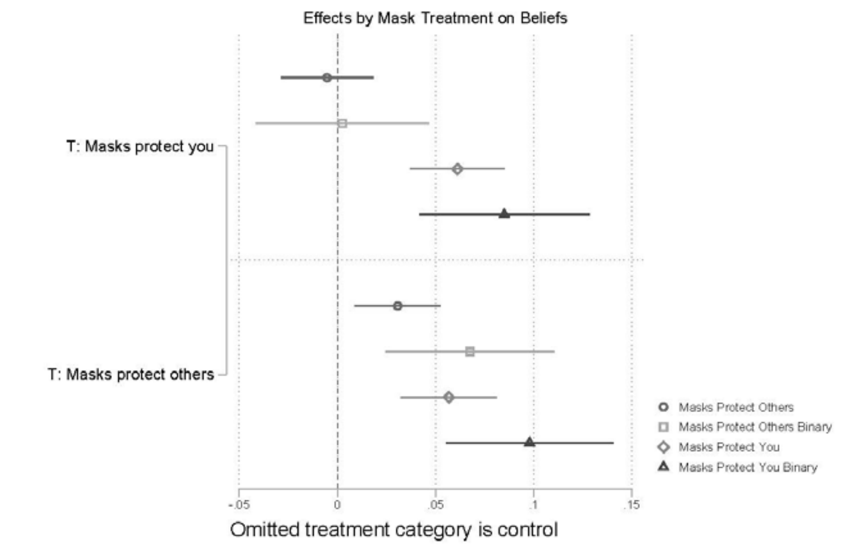 | 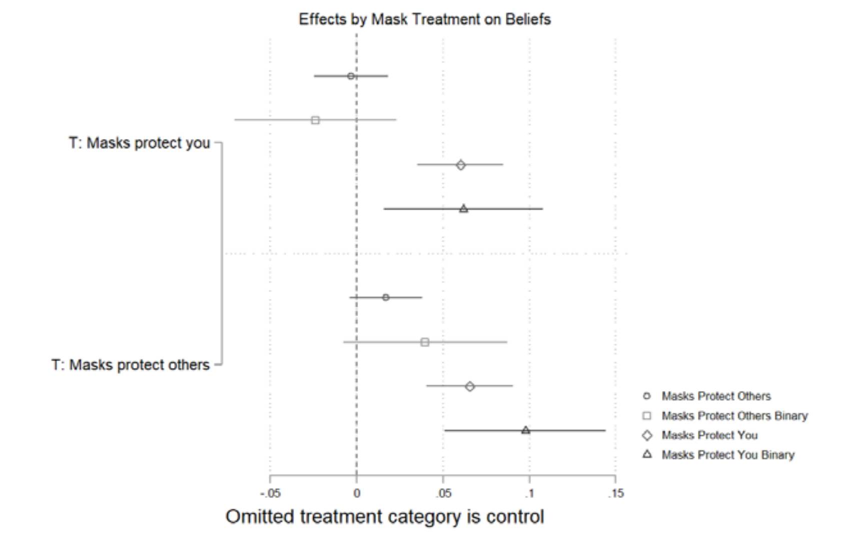 |





Effect of mask efficacy treatments on beliefs about the degree to which masks protect you and masks protect others. The figure displays OLS regression estimates with 95% confidence intervals. Models included covariates described above. For the binary outcomes, respondents who answered “strongly agree” were coded 1 and those who did not were coded 0.

**Figure S2**

| United States | Italy |
|---|---|
| 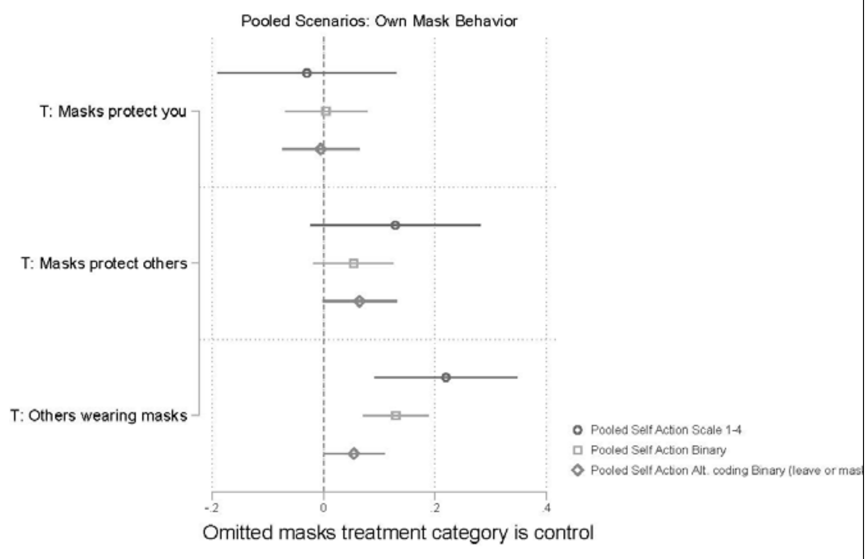 | 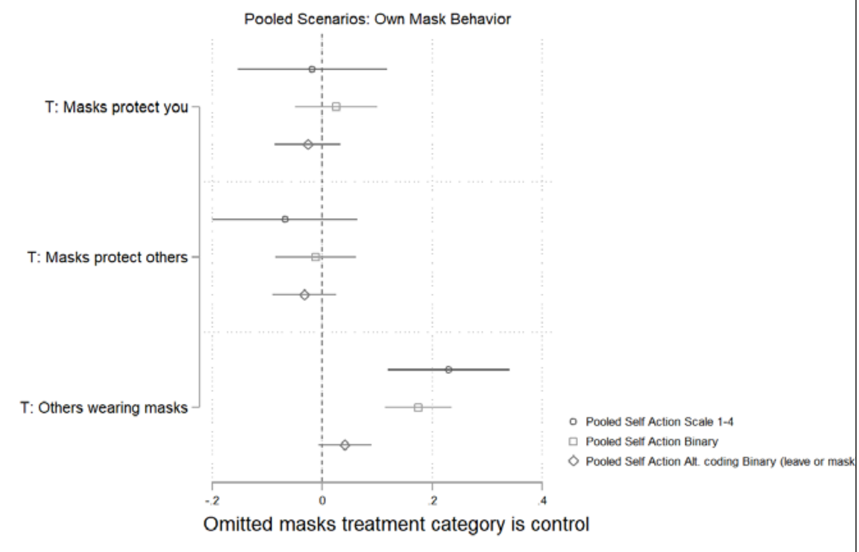 |





Effect of mask efficacy treatments and social norms treatment on reported OWN mask behavior for respondents who saw an OWN mask behavior scenario first. The figure displays OLS regression estimates with 95% confidence intervals. Models included covariates described above.

**Figure S3**

| United States | Italy |
|---|---|
| 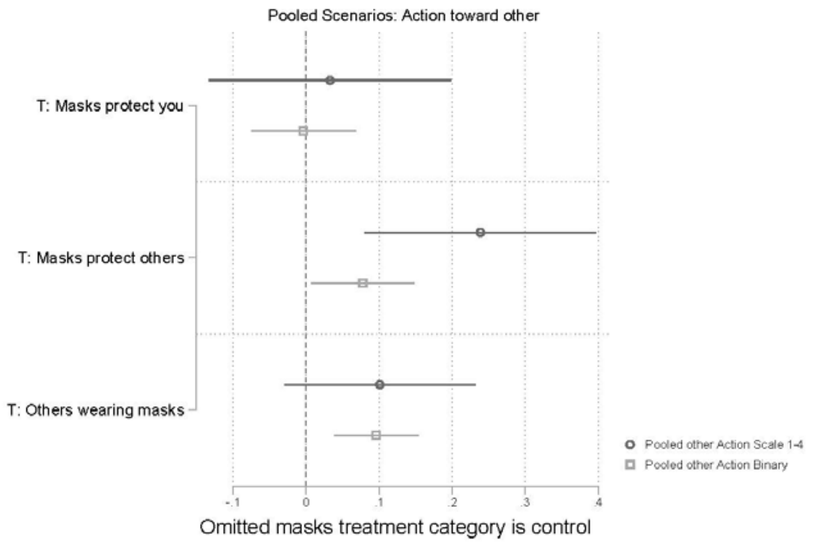 | 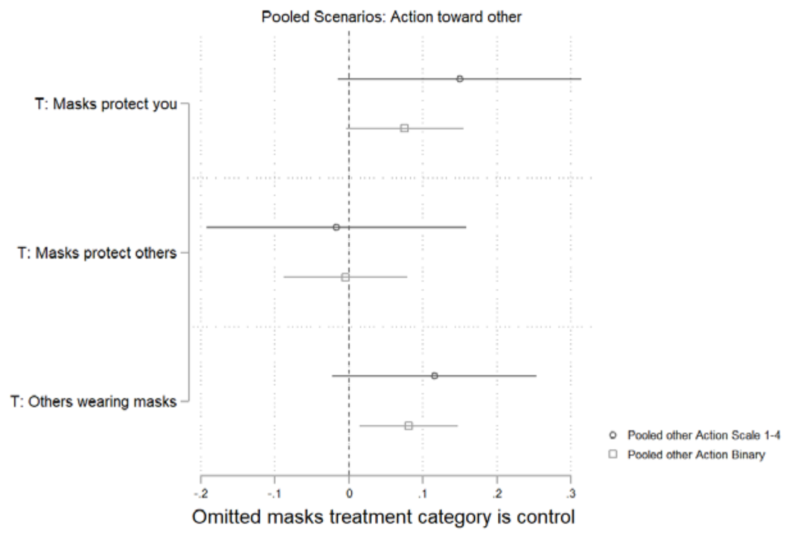 |





Effect of mask efficacy treatments and social norms treatment on reported OTHER mask behavior for respondents who saw an OTHER mask behavior scenario first. The figure displays OLS regression estimates with 95% confidence intervals. Models included covariates described above.

**Figure S4**

United States

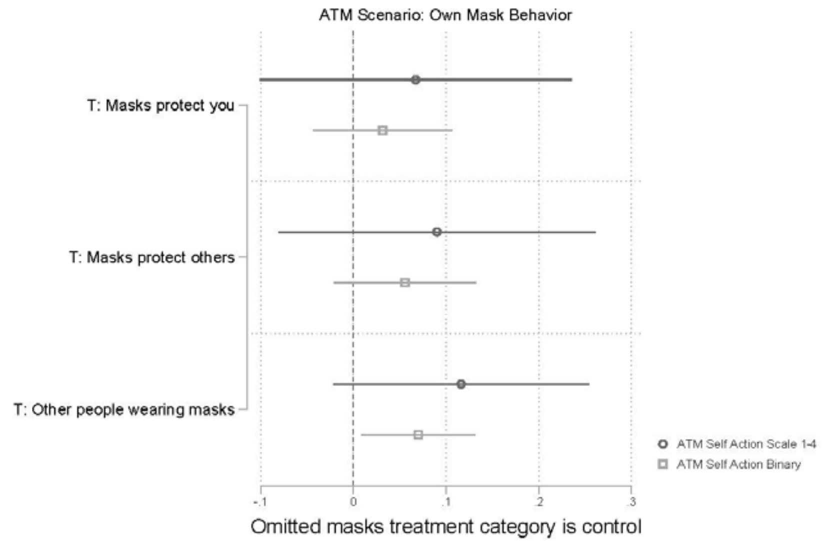


Italy

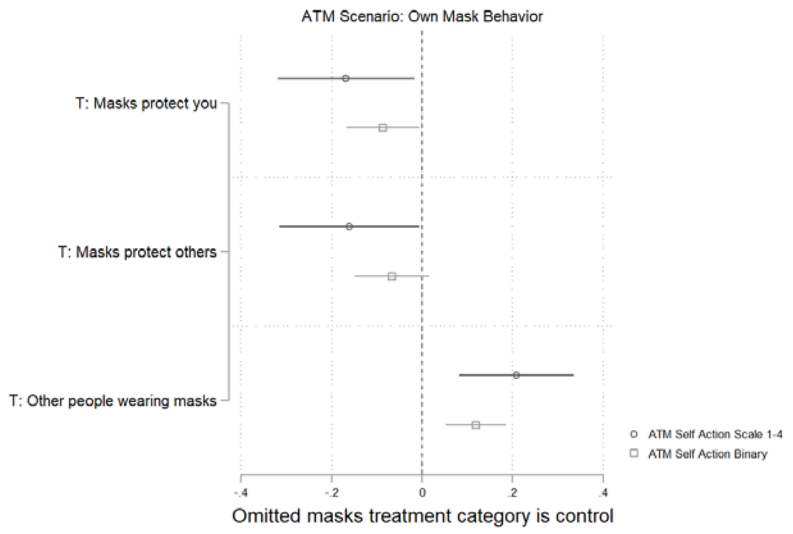


Effect of mask efficacy treatments and social norms treatment on reported OWN mask behavior for the ATM scenario. The figure displays OLS regression estimates with 95% confidence intervals. Models included covariates described above.

**Figure S5**

| United States | Italy |
|---|---|

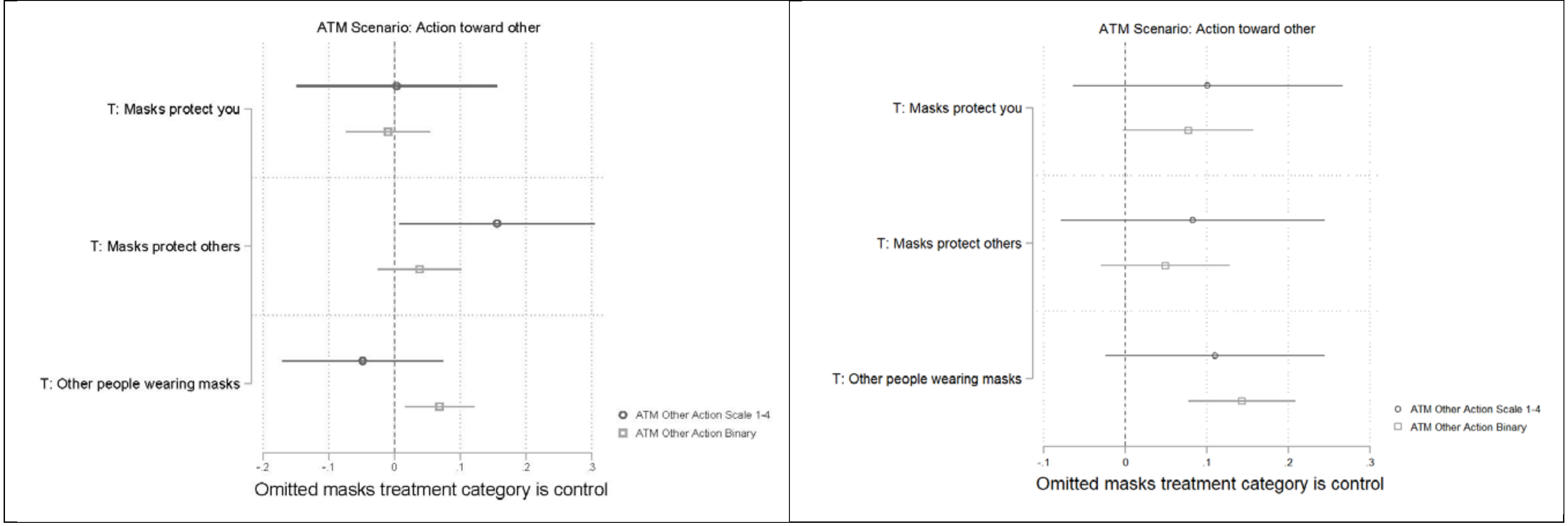


Effect of mask efficacy treatments and social norms treatment on reported action towards OTHERS for the ATM scenario. The figure displays OLS regression estimates with 95% confidence intervals. Models included covariates described above.

**Figure S6**

| United States | Italy |
| --- | --- |
| 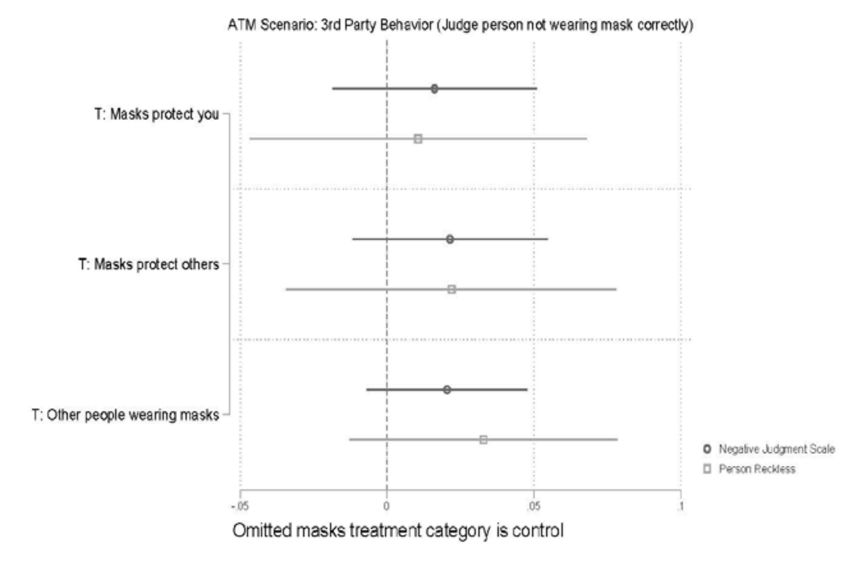 | 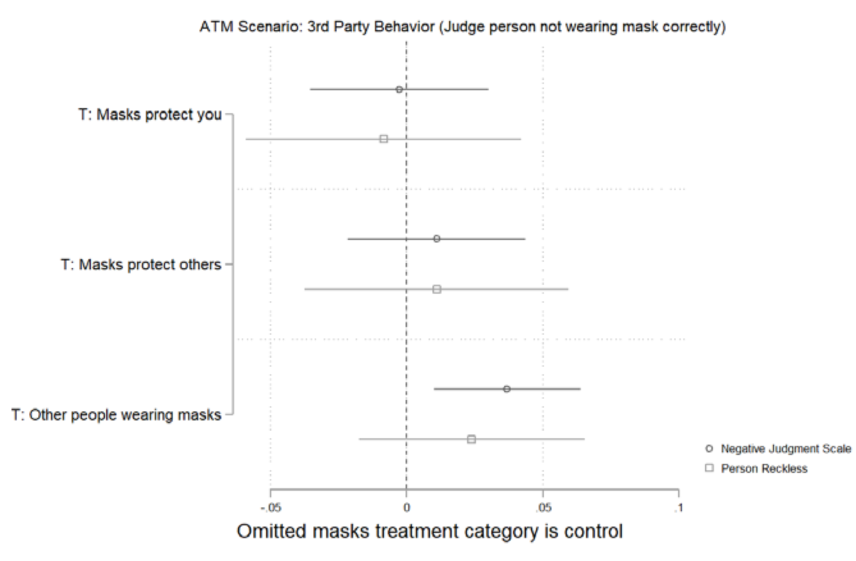 |





Effect of mask efficacy treatments and social norms treatment on reported judgment of person who is not wearing their mask correctly mask behavior for the THIRD PARTY version of ATM scenario. The figure displays OLS regression estimates with 95% confidence intervals. Models included covariates described above.

**Figure S7**

| United States | Italy |
| --- | --- |
| 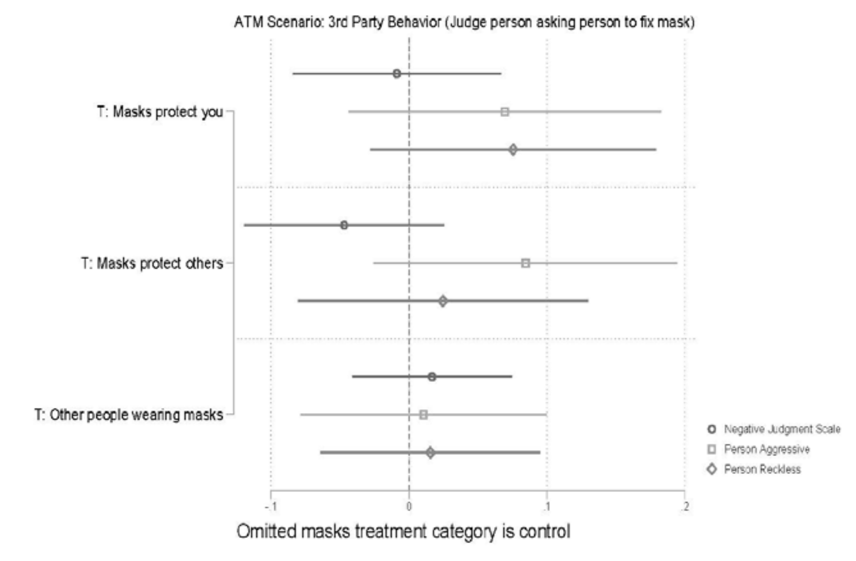 | 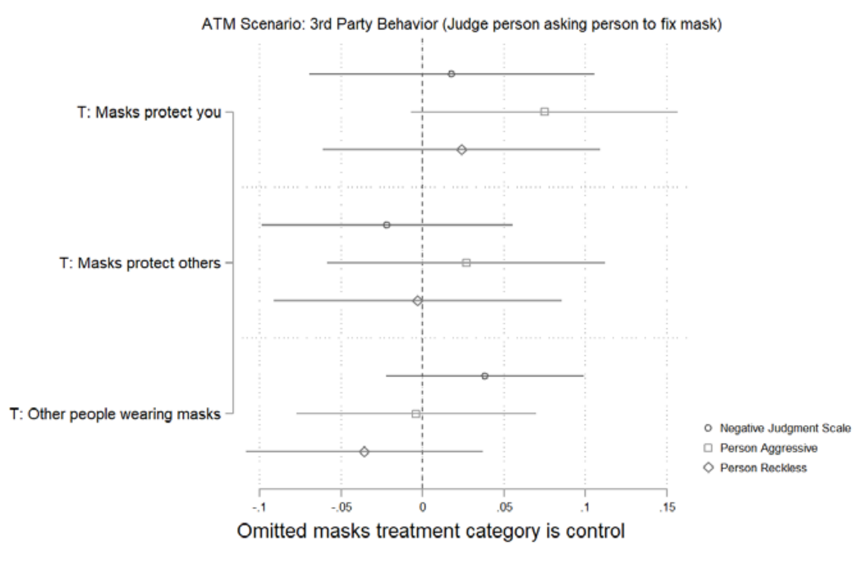 |





Effect of mask efficacy treatments and social norms treatment on reported judgment of person who asked someone to fix their mask in the THIRD PARTY version of ATM scenario. The figure displays OLS regression estimates with 95% confidence intervals. Models included covariates described above.

**Figure S8**

United States

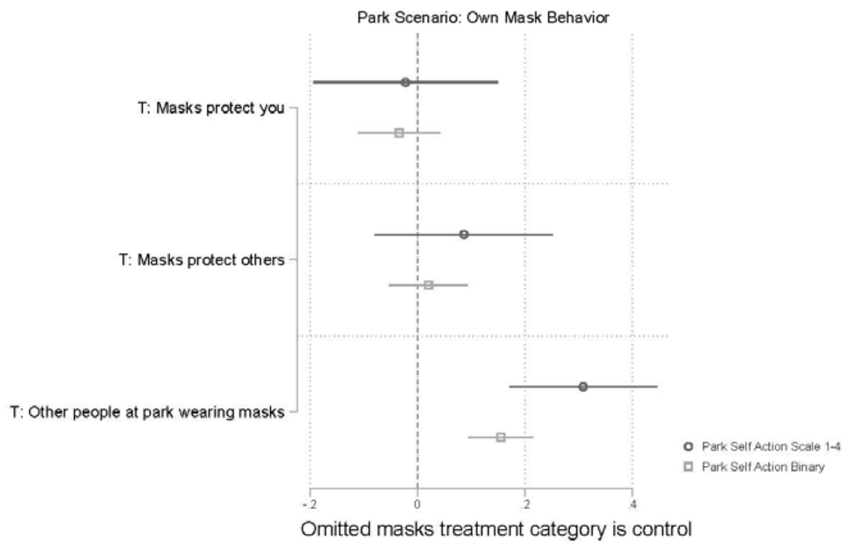


Italy

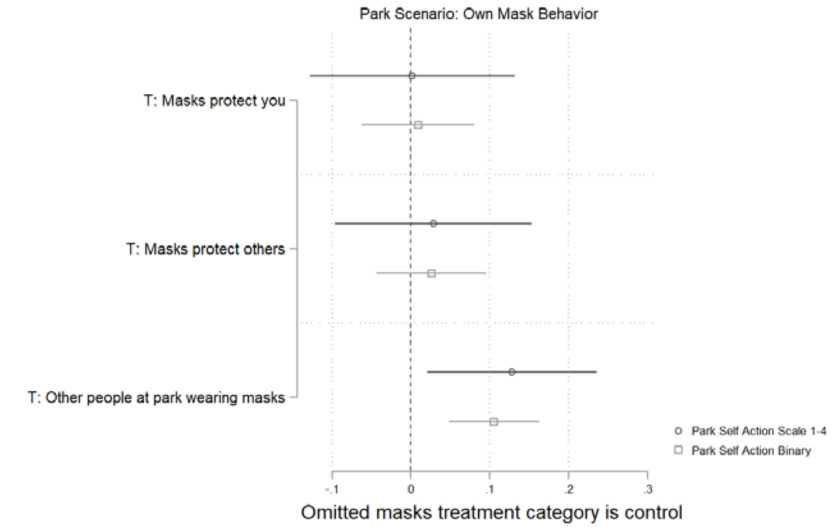


Effect of mask efficacy treatments and social norms treatment on reported OWN mask behavior for the PARK scenario. The figure displays OLS regression estimates with 95% confidence intervals. Models included covariates described above.

**Figure S9**

United States

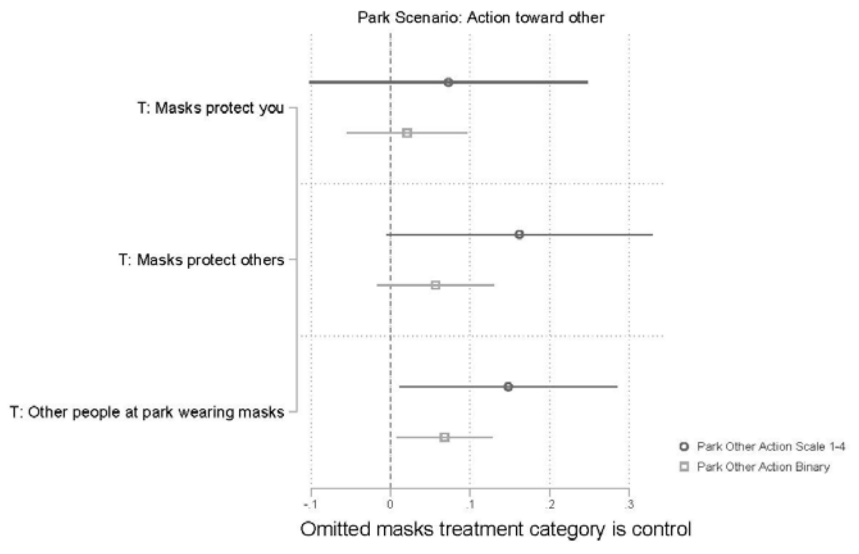


Italy

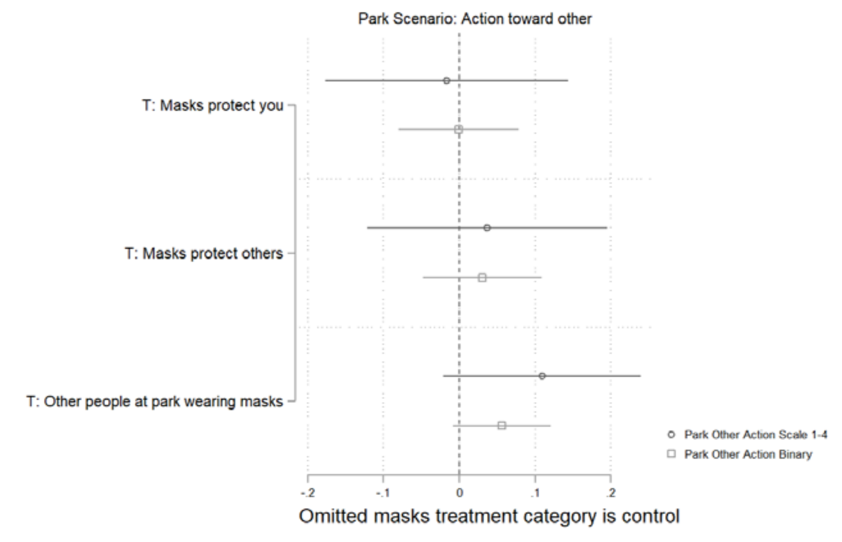


Effect of mask efficacy treatments and social norms treatment on reported action towards OTHERS for the PARK scenario. The figure displays OLS regression estimates with 95% confidence intervals. Models included covariates described above.

**Figure S10**

<table>
<tr><th>United States</th><th>Italy</th></tr>
<tr><td>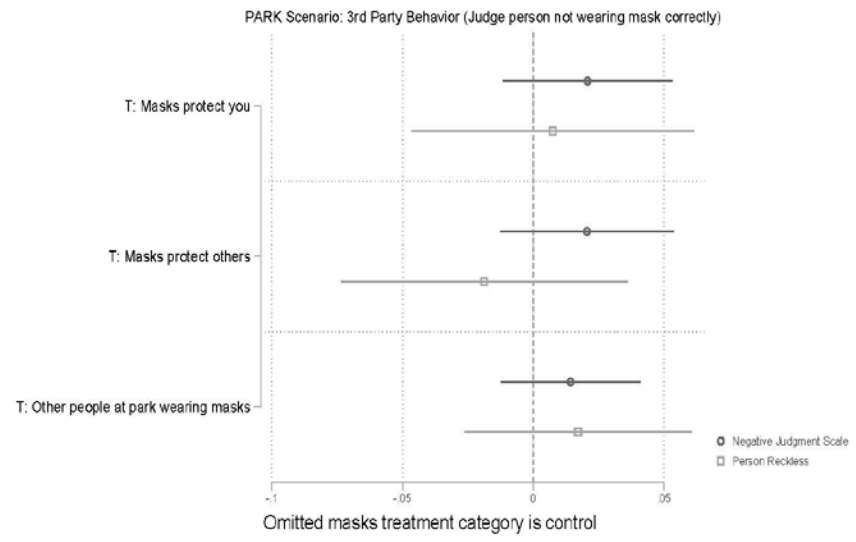
</td><td>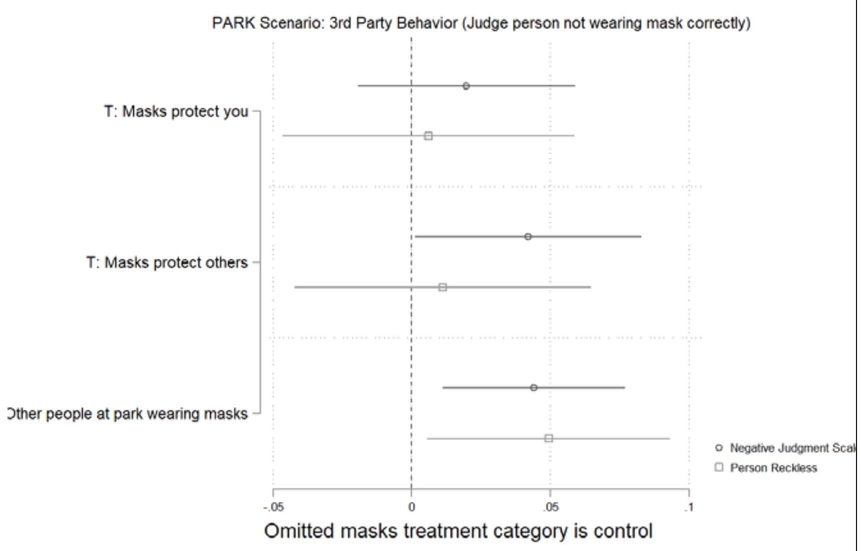
</td></tr>
</table>

Effect of mask efficacy treatments and social norms treatment on reported judgment of person who is not wearing their mask correctly mask behavior for the THIRD PARTY version of PARK scenario. The figure displays OLS regression estimates with 95% confidence intervals. Models included covariates described above.

**Figure S11**

| United States | Italy |
| --- | --- |
| 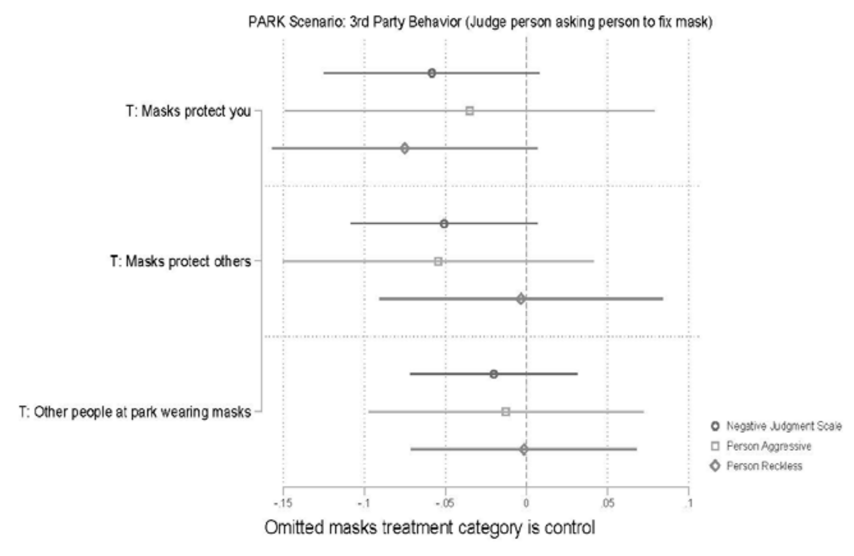 | 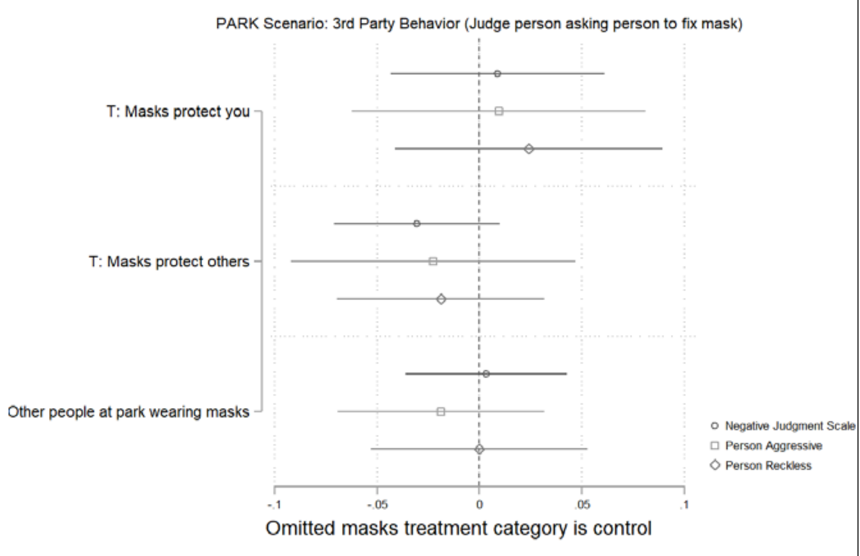 |





Effect of mask efficacy treatments and social norms treatment on reported judgment of person who asked someone to fix their mask in the THIRD PARTY version of PARK scenario. The figure displays OLS regression estimates with 95% confidence intervals. Models included covariates described above.

**Figure S12**

United States

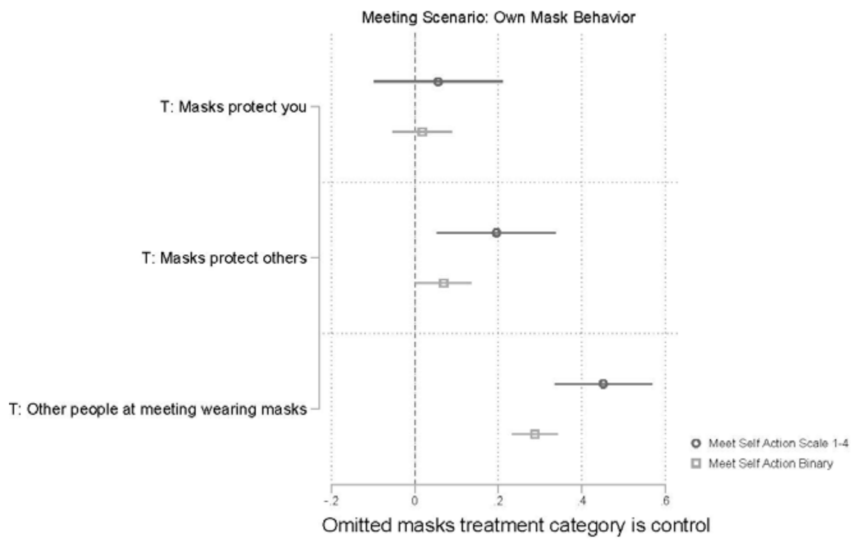


Italy

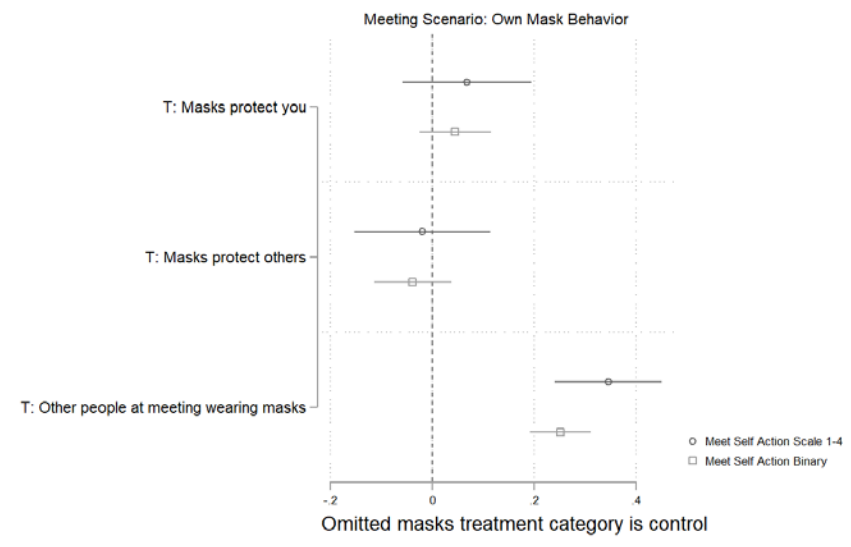


Effect of mask efficacy treatments and social norms treatment on reported OWN mask behavior for the MEETING scenario. The figure displays OLS regression estimates with 95% confidence intervals. Models included covariates described above.

**Figure S13**

United States

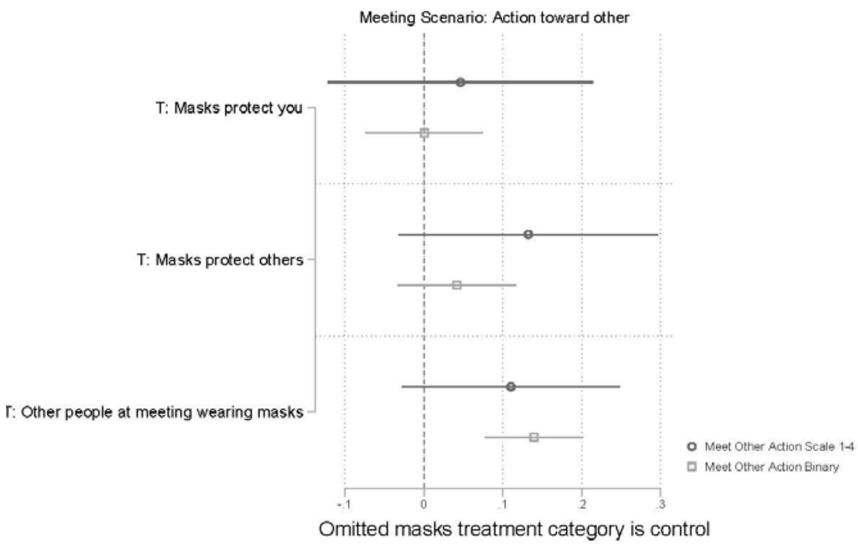


Italy

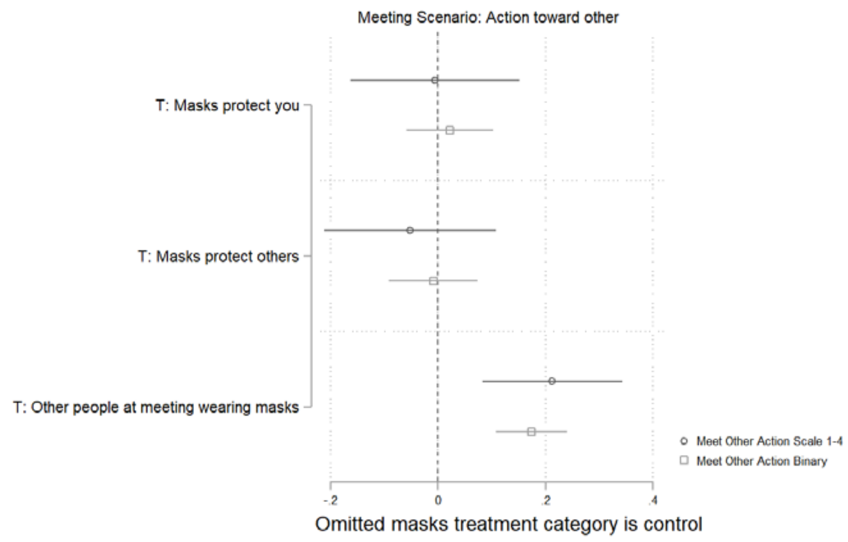


Effect of mask efficacy treatments and social norms treatment on reported action towards OTHERS for the MEETING scenario. The figure displays OLS regression estimates with 95% confidence intervals. Models included covariates described above.

**Figure S14**

United States

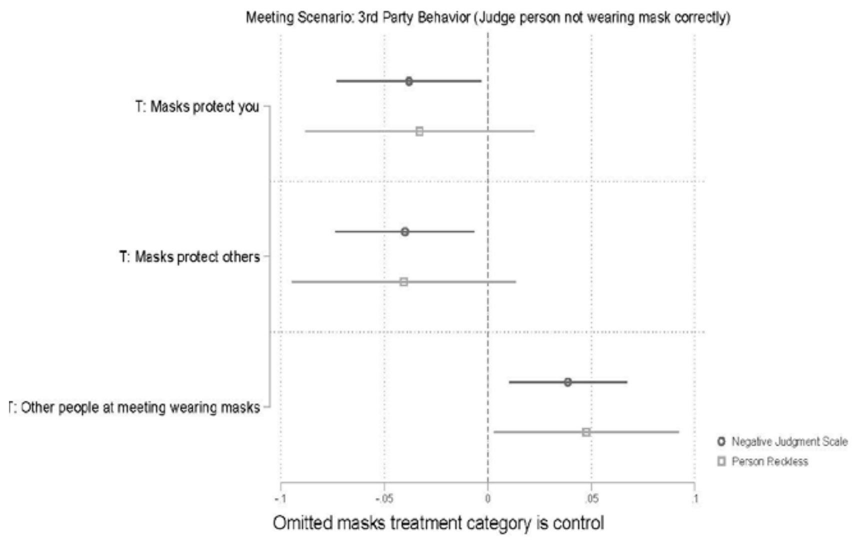


Italy

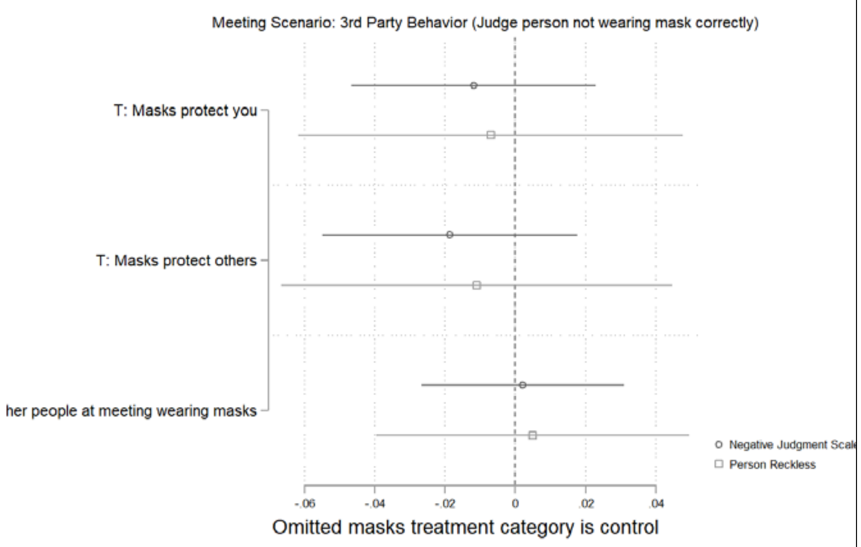


Effect of mask efficacy treatments and social norms treatment on reported judgment of person who is not wearing their mask correctly mask behavior for the THIRD PARTY version of MEETING scenario. The figure displays OLS regression estimates with 95% confidence intervals. Models included covariates described above.

**Figure S15**

| United States | Italy |
|---|---|

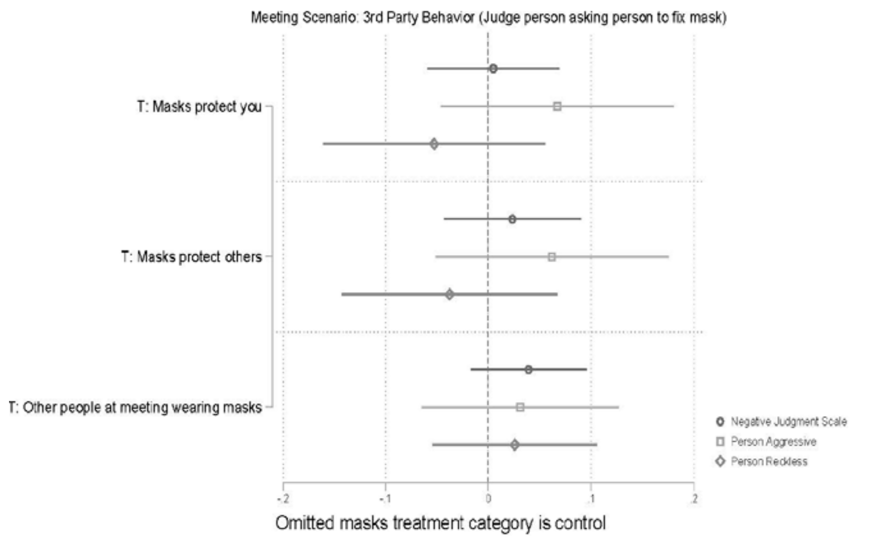


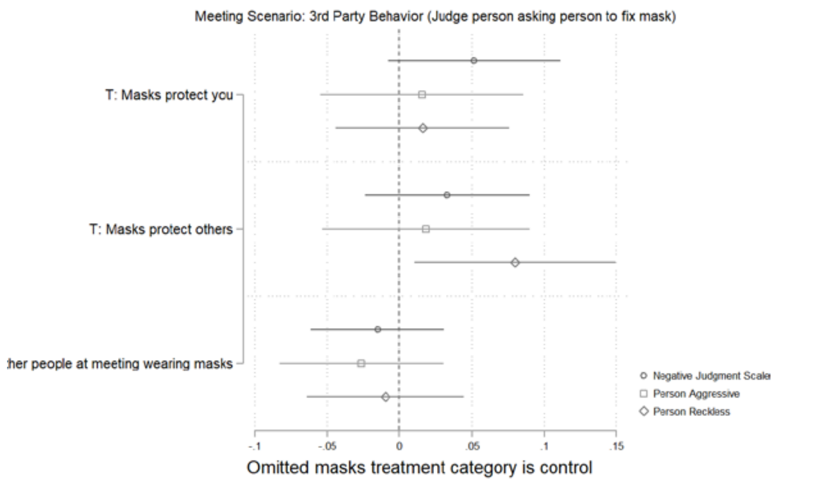


Effect of mask efficacy treatments and social norms treatment on reported judgment of person who asked someone to fix their mask in the THIRD PARTY version of MEETING scenario. The figure displays OLS regression estimates with 95% confidence intervals. Models included covariates described above.

#### S-5 Supplementary Tables

**Table S1**

| | United States | | | | Italy | | | |
|---|---|---|---|---|---|---|---|---|
| | (1) | (2) | (3) | (4) | (5) | (6) | (7) | (8) |
| | Agree masks protect others | Strongly Agree masks protect others (binary) | Agree masks protect you | Strongly Agree masks protect you (binary) | Agree masks protect others | Strongly Agree masks protect others (binary) | Agree masks protect you | Strongly Agree masks protect you (binary) |
| T: Masks protect you | -0.005 | 0.003 | 0.061 | 0.085 | -0.003 | -0.024 | 0.06 | 0.062 |
| | [0.012] | [0.022] | [0.012]*** | [0.022]*** | [0.011] | [0.024] | [0.013]*** | [0.023]*** |
| T: Masks protect others | 0.031 | 0.068 | 0.057 | 0.098 | 0.017 | 0.04 | 0.064 | 0.098 |
| | [0.011]*** | [0.022]*** | [0.012]*** | [0.022]*** | [0.011] | [0.024]* | [0.013]*** | [0.024]*** |
| Observations | 2905 | 2905 | 2904 | 2904 | 2549 | 2549 | 2550 | 2550 |
| R-squared | 0.070 | 0.067 | 0.095 | 0.090 | 0.049 | 0.036 | 0.044 | 0.026 |
| Mean of DV | 0.813 | 0.545 | 0.784 | 0.497 | 0.827 | 0.507 | 0.772 | 0.416 |
| S.D. of DV | 0.260 | 0.498 | 0.277 | 0.500 | 0.224 | 0.5 | 0.258 | 0.493 |

Corresponding OLS coefficients with robust standard errors in brackets for Figure S1. * significant at 10%; ** significant at 5%; *** significant at 1%. Placebo control condition of the mask efficacy treatment is the omitted category. Control variables and constant omitted from regression plots. Covariates are age (years), gender, household income, ethnicity (White, Black, Asian, Other), education, partisanship, work status, and previous flu vaccination. See Supplemental Information S-2 for control variable coding.

**Table S2**

| | United States | | | Italy | | |
|---|---|---|---|---|---|---|
| | (1) | (2) | (3) | (4) | (5) | (6) |
| | Pooled OWN scenario outcome | Pooled OWN behavior outcome, binary | Pooled OWN behavior outcome, alternative coding (leave or get mask), binary | Pooled OWN scenario outcome | Pooled OWN behavior outcome, binary | Pooled OWN behavior outcome, alternative coding (leave or get mask), binary |
| T: Masks protect you | 0.027 | 0.005 | 0.016 | -0.031 | -0.009 | -0.013 |
| | [0.048] | [0.022] | [0.021] | [0.040] | [0.022] | [-0.018] |
| T: Masks protect others | 0.139 | 0.057 | 0.058 | -0.052 | -0.027 | -0.016 |
| | [0.047]*** | [0.021]*** | [0.020]*** | [0.040] | [0.022] | [0.018] |
| T: Others wearing masks | 0.295 | 0.173 | 0.084 | 0.233 | 0.163 | 0.057 |
| | [0.038]*** | [0.017]*** | [0.016]*** | [0.033]*** | [0.018]*** | [0.015]*** |
| Scenario is ATM | -0.462 | -0.222 | -0.228 | -0.3 | -0.153 | -0.148 |
| | [0.049]*** | [0.022]*** | [0.021]*** | [0.042]*** | [0.022]*** | [0.019]*** |
| Scenario is MEETING | 0.077 | -0.004 | 0.058 | -0.062 | -0.054 | -0.005 |
| | [0.046] | [0.021] | [0.019]*** | [0.038]* | [0.021]*** | [0.016] |
| Observations | 2872 | 2872 | 2872 | 2530 | 2530 | 2530 |
| R-squared | 0.150 | 0.130 | 0.159 | 0.059 | 0.061 | 0.059 |
| Mean of DV | 3.111 | 0.563 | 0.656 | 3.493 | 0.694 | 0.828 |
| S.D. of DV | 1.104 | 0.496 | 0.475 | 0.842 | 0.461 | 0.377 |

Corresponding OLS coefficients with robust standard errors in brackets for Figure 1. * significant at 10%; ** significant at 5%; *** significant at 1%. Placebo control, no one/few people are wearing a mask, and the PARK scenario are the omitted categories. Control variables and constant omitted from regression plots. Covariates are age (years), gender, household income, ethnicity (White, Black, Asian, Other), education, partisanship, work status, and previous flu vaccination. See Supplemental Information S-2 for control variable coding.

**Table S3**

| | United States | | Italy | |
|---|---|---|---|---|
| | (1) | (2) | (3) | (4) |
| | Pooled OTHERS scenario outcome | Pooled OTHERS scenario outcome, binary | Pooled OTHERS scenario outcome | Pooled OTHERS scenario outcome, binary |
| T: Masks protect you | 0.037 | 0.004 | 0.026 | 0.033 |
| | [0.049] | [0.021] | [0.047] | [0.023] |
| T: Masks protect others | 0.149 | 0.045 | 0.027 | 0.026 |
| | [0.047]*** | [0.021]** | [0.047] | [0.023] |
| T: Others wearing masks | 0.060 | 0.084 | 0.143 | 0.125 |
| | [0.039] | [0.017]*** | [0.039]*** | [0.019]*** |
| Scenario is ATM | -0.262 | -0.212 | -0.433 | -0.239 |
| | [0.047]*** | [0.021]*** | [0.047]*** | [0.023]*** |
| Scenario is MEETING | -0.079 | -0.105 | -0.054 | -0.055 |
| | [0.049] | [0.022]*** | [0.047] | [0.023] |
| Observations | 2868 | 2868 | 2530 | 2530 |
| R-squared | 0.086 | 0.085 | 0.086 | 0.091 |
| Mean of DV | 2.724 | 0.343 | 3.176 | 0.557 |
| S.D. of DV | 1.073 | 0.475 | 1.002 | 0.497 |

Corresponding OLS coefficients with robust standard errors in brackets for Figure 2. * significant at 10%; ** significant at 5%; *** significant at 1%. Placebo control, no one/few people are wearing a mask, and the PARK scenario are the omitted categories. Control variables and constant omitted from regression plots. Covariates are age (years), gender, household income, ethnicity (White, Black, Asian, Other), education, partisanship, work status, and previous flu vaccination. See Supplemental Information S-2 for control variable coding.

**Table S4**

| | United States | | Italy | |
|---|---|---|---|---|
| | (1) Negative judgment person not wearing mask correctly | (2) Negative judgment person asking other to fix mask | (3) Negative judgment person not wearing mask correctly | (4) Negative judgment person asking other to fix mask |
| T: Masks protect you | 0.001 | -0.022 | -0.002 | 0.028 |
| | [0.010] | [0.019] | [0.01] | [0.019] |
| T: Masks protect others | 0.001 | -0.022 | 0.005 | 0.005 |
| | [0.010] | [0.019] | [0.01] | [0.017] |
| T: Others wearing masks | 0.015 | -0.036 | 0.02 | 0.007 |
| | [0.008]* | [0.016]** | [0.009]** | [0.015] |
| Scenario is ATM | -0.011 | 0.046 | -0.057 | 0.004 |
| | [0.010] | [0.019]** | [0.01]*** | [0.018] |
| Scenario is MEETING | -0.053 | 0.006 | -0.096 | 0.042 |
| | [0.010]*** | [0.018] | [0.011]*** | [0.016]** |
| Observations | 2865 | 686 | 2172 | 555 |
| R-squared | 0.101 | 0.115 | 0.072 | 0.094 |
| Mean of DV | 0.616 | 0.259 | 0.806 | 0.103 |
| S.D. of DV | 0.226 | 0.208 | 0.205 | 0.177 |

Corresponding OLS coefficients with robust standard errors in brackets for Figure 3. * significant at 10%; ** significant at 5%; *** significant at 1%. Placebo control, no one/few people are wearing a mask, and the PARK scenario are the omitted categories. Control variables and constant omitted from regression plots. Covariates are age (years), gender, household income, ethnicity (White, Black, Asian, Other), education, partisanship, work status, and previous flu vaccination. See Supplemental Information S-2 for control variable coding.

**Table S5**

| | United States | | Italy | |
|---|---|---|---|---|
| | Mean | SD | Mean | SD |
| Age | 45.2 | 17.3 | 46.8 | 15 |
| Proportion Female | 0.482 | 0.5 | 0.526 | 0.5 |
| Proportion Democrat | 0.465 | 0.499 | | |
| Proportion Republican | 0.389 | 0.487 | | |
| Proportion Left coalition | | | 0.177 | 0.381 |
| Proportion Right coalition | | | 0.255 | 0.436 |
| Proportion 5 Star Movement | | | 0.247 | 0.431 |
| Proportion College Education | 0.433 | 0.496 | 0.218 | 0.413 |
| Proportion White | 0.725 | 0.447 | | |
| Proportion Black | 0.125 | 0.330 | | |
| Proportion Northeast United States | 0.203 | 0.402 | | |
| Proportion Midwest United States | 0.188 | 0.391 | | |
| Proportion Southern United States | 0.377 | 0.485 | | |
| Proportion Northeast Italy | | | 0.189 | 0.392 |
| Proportion Northwest Italy | | | 0.269 | 0.443 |
| Proportion Center Italy | | | 0.2 | 0.4 |
| Proportion South Italy | | | 0.243 | 0.429 |
| Proportion Islands Italy | | | 0.099 | 0.298 |

Descriptive statistics for United States and Italy experiments.

#### S-6 References


1. D. K. Chu *et al.*, Physical distancing, face masks, and eye protection to prevent person-to-person transmission of SARS-CoV-2 and COVID-19: a systematic review and meta-analysis. *Lancet* **395**, 1973-1987 (2020).
2. M. Mills, C. Rahal, E. Akimova (Face masks and coverings for the general public: behavioural knowledge, effectiveness of cloth coverings and public messaging [Internet]. The Royal Society & The British Academy; 2020.
3. E. E. Sickbert-Bennett *et al.*, Filtration Efficiency of Hospital Face Mask Alternatives Available for Use During the COVID-19 Pandemic. *JAMA Intern Med* 10.1001/jamainternmed.2020.4221 (2020).
4. A. Konda *et al.*, Aerosol Filtration Efficiency of Common Fabrics Used in Respiratory Cloth Masks. *ACS Nano* **14**, 6339-6347 (2020).
